# Life course exposures continually shape antibody profile and risk of seroconversion to influenza

**DOI:** 10.1101/2020.01.15.19015693

**Authors:** Bingyi Yang, Justin Lessler, Huachen Zhu, Chao Qiang Jiang, Jonathan M. Read, James A. Hay, Kin On Kwok, Ruiyin Shen, Yi Guan, Steven Riley, Derek A.T. Cummings

## Abstract

Complex exposure histories and immune mediated interactions between influenza strains contribute to the life course of human immunity to influenza. Antibody profiles can be generated by characterizing immune responses to multiple antigenically variant strains, but how these profiles vary across individuals and determine future responses is unclear. We used hemagglutination inhibition titers from 21 H3N2 strains to construct 777 paired antibody profiles from people aged 2 to 86, and developed novel metrics to capture features of these profiles. Total antibody titer per potential influenza exposure increases in early life, then decreases in middle age. Increased titers to one or more strains were seen in 97.8% of participants, suggesting widespread influenza exposure. While titer changes were seen to all strains, recently circulating strains exhibited the greatest variation. Higher pre-existing, homologous titers reduced the risk of seroconversion to recent strains. After adjusting for homologous titer, we also found an increased frequency of seroconversion among those with higher immunity to older previously exposed strains. Our results suggest that a comprehensive quantitative description of immunity encompassing past exposures could lead to improved correlates of risk of influenza.

## Main Text

Seasonal influenza remains a ubiquitous threat to human health. It is estimated to kill between 291,000 and 645,000 people each year worldwide (*1*). Through the process of antigenic drift, antigenically novel strains replace previously circulating viruses every few influenza seasons (*2*). As a result, people can be infected multiple times over a lifetime (*3*). Each of these infections leaves a mark on a person’s immune system, and the accumulation of antibody responses over a life course leads to complex individual antibody profiles reflecting both recent and past exposures (*3*–*6*). A growing body of evidence suggests that the order and timing of influenza exposures shape the immune response in ways that may affect morbidity and mortality (*3, 7*), particularly when encountering novel (i.e. pandemic or potentially pandemic) strains (*8*), yet a comprehensive quantitative description of how past exposure to multiple strains shapes infection risk remains elusive.

Historically, most studies have focused on measuring antibodies to a single strain or small set of strains of a subtype, either to measure the incidence of influenza through changes in titers (*9*–*11*) or as a proxy for immunity (e.g., as an endpoint in vaccine efficacy studies) (*12*–*15*). Exceptions do exist, and studies of multi-strain dynamics after immunological challenge date back to at least 1941 (*16*). However, historically, few analytical approaches were available with which to interpret these complex data. In recent years there has been increasing interest in measuring and interpreting the breadth and strength of antigenic profiles across a diverse set of influenza strains (*4, 7*). It is believed to provide a more nuanced picture of the humoral immune response to influenza than measuring titers to single strains (i.e. homologous titers), while the quantitative role of past exposures in determining risk of future infections and subsequent immune responses is still unclear.

Here, we describe paired antibody profiles measured at two time points (baseline from 2009 to 2011 and follow-up from 2014 to 2015), roughly four years apart, in a large sample of individuals from an ongoing cohort study in Guangzhou, Guangdong Province, China (*17*). We measured immune responses to multiple chronologically ordered H3N2 influenza strains (referred to as antibody profiles) that represent the history of H3N2 circulation in humans since its emergence in 1968. We aim to determine how those profiles vary across individuals and between study visits, and to test if there exist features of these antibody profiles that are more predictive of the risk of seroconversion to recently circulating strains than homologous titers only.

Antibody responses to 21 H3N2 influenza strains were measured for each individual from sera using hemagglutination inhibition assays (HAI). The strains selected covered the period between 1968-2014, and included strains isolated at 2-3 year intervals. Strains included in the vaccine formulation and tested by Fonville et al. (*7*) were prioritized, but when this was not possible another antigenically representative strain from that year was selected. We described individual antibody profiles from these serological data, and introduced multiple novel metrics to summarize each individual’s profile and facilitate comparisons of profiles across age and time. We also determined if summary metrics of antibody profiles improved predictions of the risk of seroconversion (i.e. four-fold or greater rise), a commonly used indicator of influenza exposure (*14*).

### Serological data and individual antibody profiles

Both baseline and follow-up sera were available from 777 participants aged from 2 to 86 years old (age at baseline is used throughout unless otherwise noted) (*17*). Participants giving serum were largely representative of the censused and overall study populations, with under representation of young children and over representation of participants aged 40-59 years (table S1) (*17*). There were 10 participants (1.3%) who reported being vaccinated against influenza at baseline, and 5 participants (0.6%) who reported influenza vaccination between the two visits.

In total, 32,606 HAI titer readings were available, with only 28 missing out of 21 × 777 × 2 possible strain-individual-visit combinations. Fig. 1 shows example individual antibody profiles for an age representative subset of individuals. We defined the four strains (i.e. A/Perth/2009, A/Victoria/2009, A/Texas/2012 and A/HongKong/2014) that were isolated between the first baseline and last follow-up as “recent strains”. Strains isolated prior to an individual’s birth were defined as “pre-birth strains” (Fig. 1, A and C) and “post-birth strains” otherwise.

**Fig. 1.**
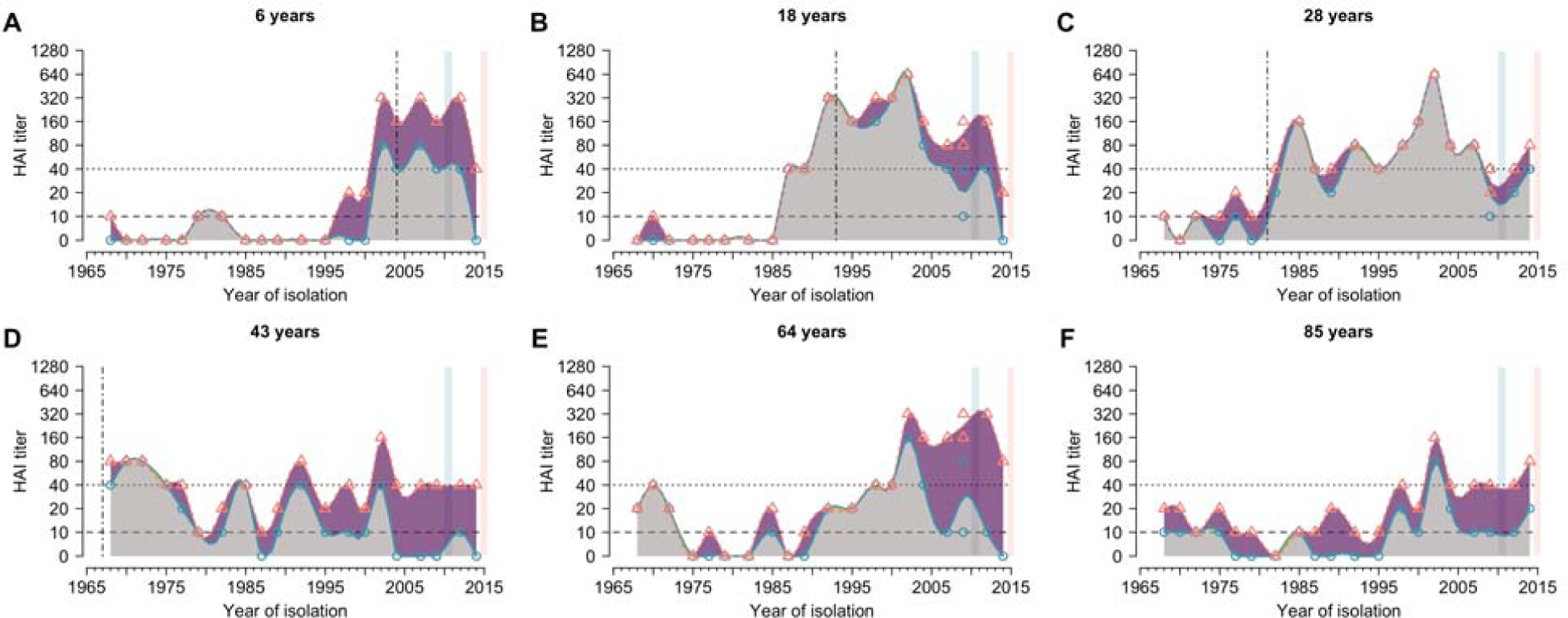
Representative individual profiles of HAI titer against H3N2 strains circulating over forty years. Blue circles and red triangles represent the HAI titers against the tested strains at baseline and follow-up visit, respectively. Blue and red solid lines represent the smoothed HAI titers for serum collected from baseline and follow-up visit, respectively. Smooth splines of HAI titers on circulating years are shown in this figure for illustration purposes and not used in the subsequent analysis. Grey areas represent the baseline antibody profile. Purple and green areas indicate the increase and decrease of HAI titer at follow-up visit compared to baseline, respectively. Blue and red vertical blocks represent the duration for baseline and follow-up visit, respectively. Vertical dotted-dashed lines indicate the year of birth of the individual. Dashed and dotted lines represent the titer of 1:10 (detectable cutoff) and 1:40 (protective cutoff), respectively.

All individuals had detectable titers (1:10) and 95.6% had protective titers (1:40) to at least one strain at baseline (Fig 2A-B). The median of geometric mean titer (GMT) across all 21 tested strains was 14.3 [interquartile range (IQR), 11.4 to 25.9) at baseline and 36.4 (IQR, 21.9 to 46.8) at follow-up (Fig. 2, A and B, and table S2). GMTs of post-birth strains [18.3; 95% confidence interval (CI), 18.0 to 18.7] were higher than that of pre-birth strains (9.6; 95% CI, 9.2 to 10.0) at baseline (Fig. 2A). Participants had the highest titer to strains that were isolated within the first decade of their life (4.3 years; IQR, 2.0 to 6.9 years across strains) (fig. S1). GMTs of recent strains (12.0; 95% CI, 11.6 to 12.4) were lower than that of non-recent strains (18.7; 95% CI, 18.3 to 19.0) at baseline, but were higher at follow-up [41.5 (95% CI, 39.7 to 43.3) for recent strains and 31.3 (95% CI, 30.7 to 31.9) for non-recent strains].

**Fig. 2.**
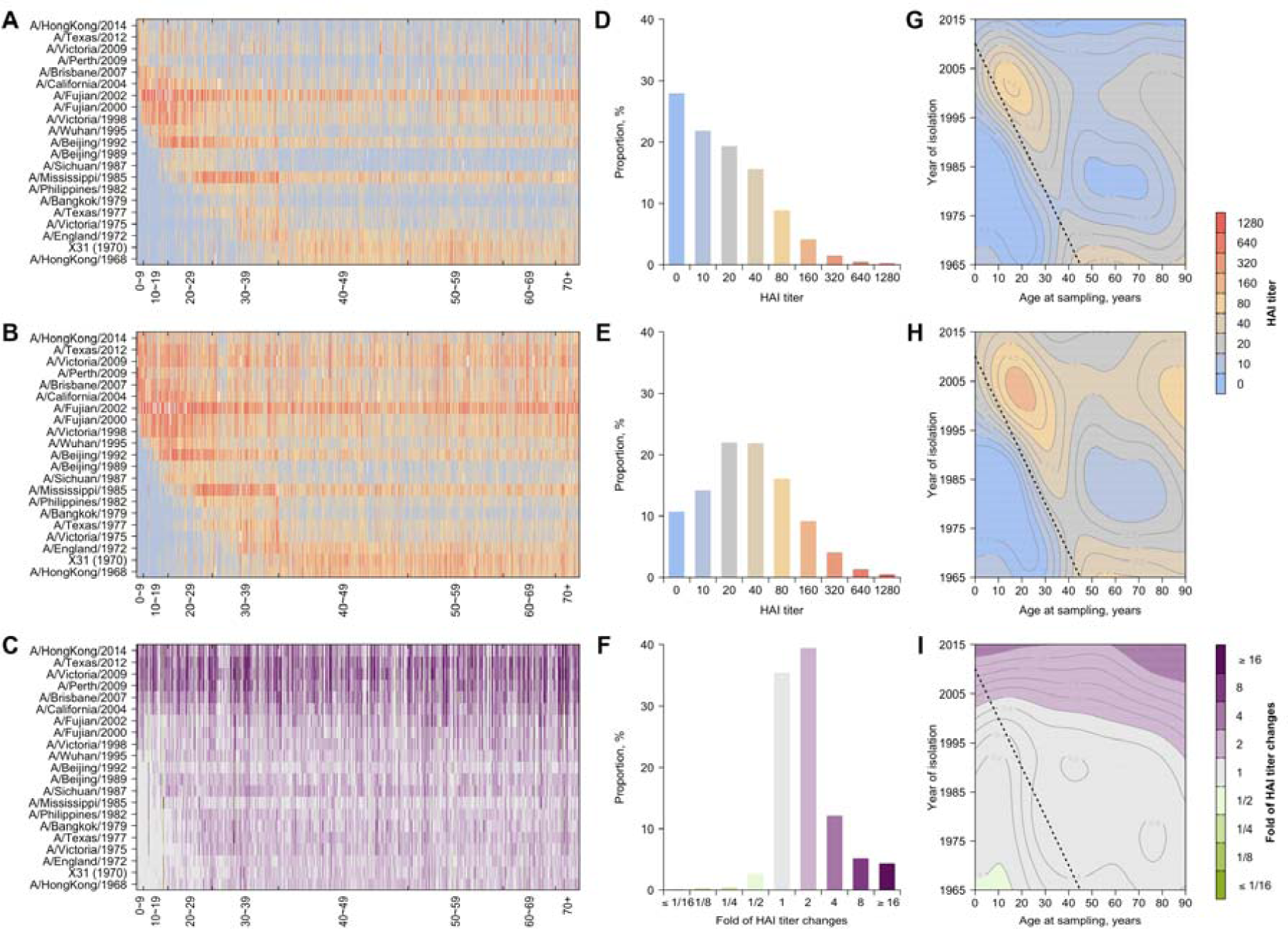
HAI titers and differences in HAI titers between two visits against historical H3N2 strains. Panel **A** and **B**: HAI titers against H3N2 strains from serum collected from baseline and follow-up visit, respectively. Participants are sorted by age at baseline sampling. Panel **C**: Fold of changes of HAI titers between two visits. Cell of row *i* and column *j* represents HAI titer or differences of HAI between two visits to strain *i* for person *j*. Panel D-F: Distribution of HAI titers from serum collected from baseline (**D**), follow-up visit (**E**), and fold of changes between two visits (**F**), respectively. Distributions were plotted after grouping titers across all participants and all tested H3N2 strains. (**G**) Variations of HAI titers at baseline with ages of participants and the year of isolation of tested strains. Dashed lines represent birth years of the participants (same for panel H and I). (**H**) Variations of HAI titers at follow-up with ages of participants and the year of isolation of tested strains. (**I**) Variation of changes in HAI titers between the two visits with ages of participants and the year of isolation of tested strains.

### Summary metrics of antibody profiles

We hypothesized that features of an antibody profile determine an individual’s risk of seroconversion over and above homologous titer. We developed several metrics aimed at summarizing the information in individuals’, often complex, antibody profiles (Fig. 3). We estimated: the *area under the curve* (AUC) for each antibody profile (i.e. the integral of an individual’s measured log titers); the *width* (W_z_) of an individual’s antibody titer above a threshold *z* (i.e. the proportion of the profile above that threshold; W_40_ for protective threshold and W_10_ for detectable threshold); and the *average titer year* (ATY) of each antibody profile (i.e. the average of strain isolation years weighted by their titer) (see Methods). We hypothesized that these features of antibody profiles captured biologically relevant properties of the immune response to H3N2; in particular, overall levels of antibody mediated immunity (for AUC), the breadth of antibody mediated immune response (for W_40_ and W_10_) and temporal center of mass of H3N2 immunity (for ATY). In most analyses, we use normalized versions of these metrics (i.e. nAUC, nW_40_, nW_10_, nATY) to adjust for differences between individuals in the number of possibly exposed strains given their ages (i.e. individuals could not have been exposed to pre-birth strains) (see Methods. Non-normalized analysis included in Supplementary Materials, fig. S2, tables S3 and S4).

**Fig. 3.**
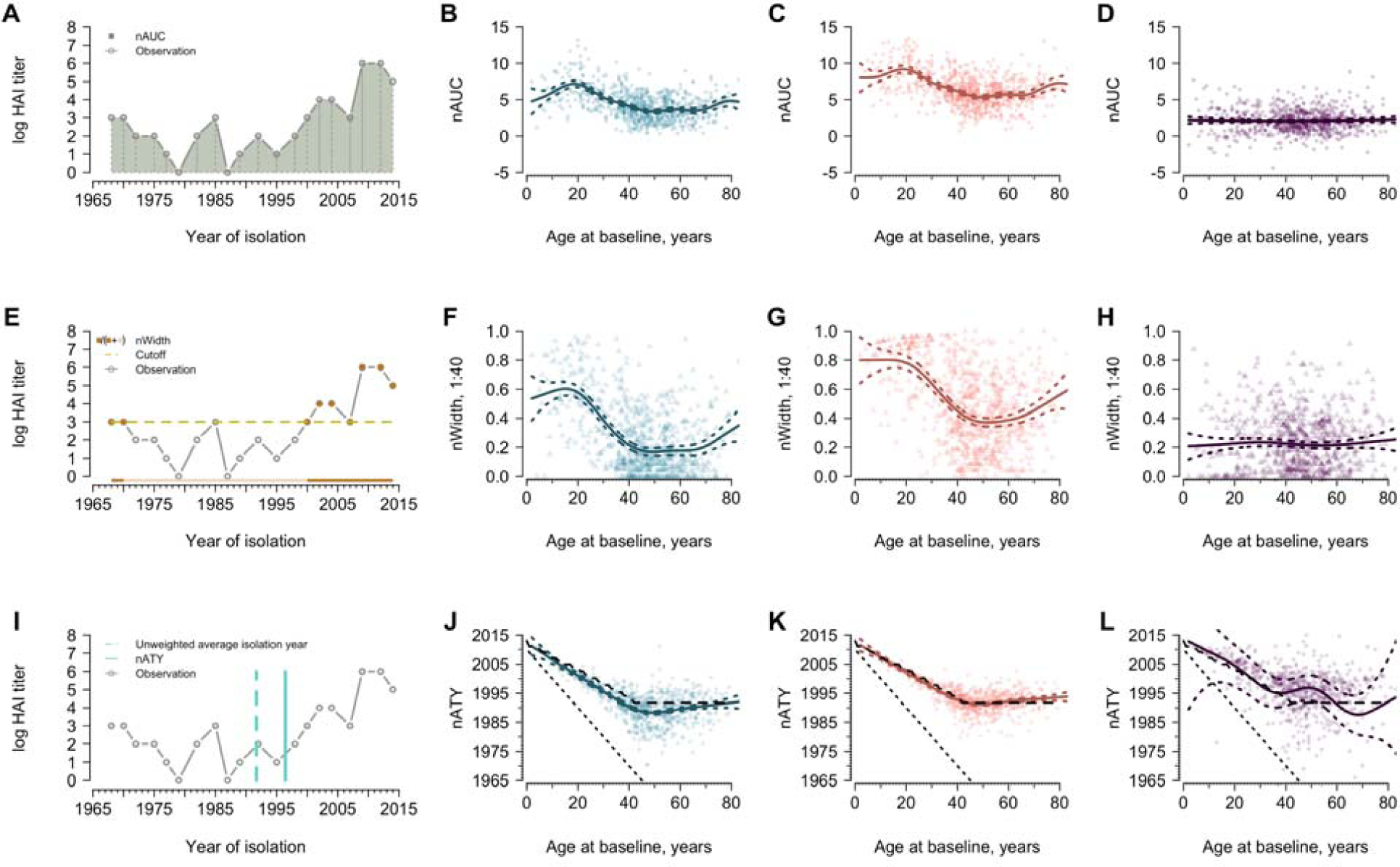
The normalized area under the curve (nAUC), width above 1:40 (nW_40_) and average titer years (nATY) varying with age. Metrics were calculated using post-birth strains and normalized by the number of post-birth strains. Blue and red represent the metrics measured for serum collected from baseline and follow-up visit, respectively. Purple indicates the differences in metrics between the two visits. Solid lines are predictions from a generalized additive model and the colored dashed lines represent the corresponding 95% confidence intervals. (**A**) Demonstration of nAUC for one participant as an example. The same participant is used for panel E and I. (**B**) Age and nAUC at baseline. (**C**) Age and nAUC at follow-up. (**D**) Age and changes in nAUC between the two visits. (**E**) Demonstration of nW_40_ for one participant as an example. Solid points are the years that contributed to calculating width. (**F**) Age and nW_40_ at baseline. (**G**) Age and nW_40_ at follow-up. (**H**) Age and changes in nW40 between the visits. (**I**) Demonstration of nATY for one participant as an example. (**J**) Age and nATY at baseline. The sloping black dotted lines indicate the year of birth of participants and the dashed lines indicate the unweighted average isolation year of post-birth strains given age on x-axis (same for panel K and L). (**K**) Age and nATY at follow-up. (**L**) Age and changes in nATY between the visits.

We calculated nAUC, nW40, nW10, nATY for each participant, and fitted generalized additive models using a spline on age to estimate the association of these metrics with participant age. The nAUC of titers increased with participant age, peaking at ∼ 20 years of age for both visits and gradually decreasing to a low at ∼ 50 years of age (Fig. 3, B and C). The average nAUC among participants aged 20 years old was estimated to be higher than that among participants aged 50 years old [ratio: 2.1 (95% CI, 1.9 to 2.4) for baseline and 1.7 (95% CI, 1.5 to 1.9) for follow-up]. After the age of 70, nAUC started to increase.

The width of HAI titers above protective titer levels (nW_40_) illustrated an increasing trend with age grows until approximately 15 years of age at the time of sample collection, peaking at 60.3% and 80.1% at baseline and follow-up, respectively (Fig. 3, F and G). There was a more distinct drop in nW_40_ among participants aged 50 years old [ratio to peak: 0.28 (95% CI, 0.25 to 0.29) and 0.46 (95% CI, 0.43 to 0.49) for baseline and follow-up, respectively], compared to the drop in nW_10_ [ratio to peak: 0.78 (95% CI, 0.75 to 0.79) and 0.94 (95% CI, 0.90 to 0.97)] (fig. S3). An uptick of widths among those older than 60 years was observed for both cutoffs (Fig. 3 and fig. S**3**).

The intent of nATY is to help us understand how immunity to strains circulating at different times contribute to the overall immune profile. At the extremes, if the antibody response was completely dominated by early infections, nATY would track with birth year, while if all strains circulating in one’s life were equally important, nATY would track with the midpoint of the time at risk (i.e. unweighted average isolation year of post-birth strains). Our empirical observations are more in line with the latter hypothesis; baseline nATY moved away from birth year at a rate of 0.50 years for every additional year of life (0.45 years at follow-up) (Fig. 3, J and K).

### Changes in antibody profiles between the two visits

Across our cohort, changes, particularly increases, in titer were seen across all strains, including strains that have long been extinct (Fig. 2 C and F and Fig. 4A). 97.9% of people experienced a rise against one or more strains. 73.7% showed a 4-fold or greater titer increase (seroconversion) to one or more (Fig. 2I), with increased risk of occurring in strains circulating after 1998 compared to A/HongKong/1968 [odds ratio (OR) ranges from 2.7 (95% CI, 2.0 to 3.9) to 16.4 (95% CI, 12.0 to 22.6) across these strains] (fig. S4), i.e. strains more antigenically similar to ones with which the participant could have been infected;. The largest increases in HAI titers were detected for four recent strains (Fig. 4B), and there was high-correlation in seroconversions; 63.1% of those who seroconverted to one or more strains seroconverted to all four strains. (fig. S5). Meanwhile, a minority of people (18.3%) showed a decrease in titer to one or more strains, likely due to transient boosting from exposures prior to baseline sampling (*5, 18*).

**Fig. 4.**
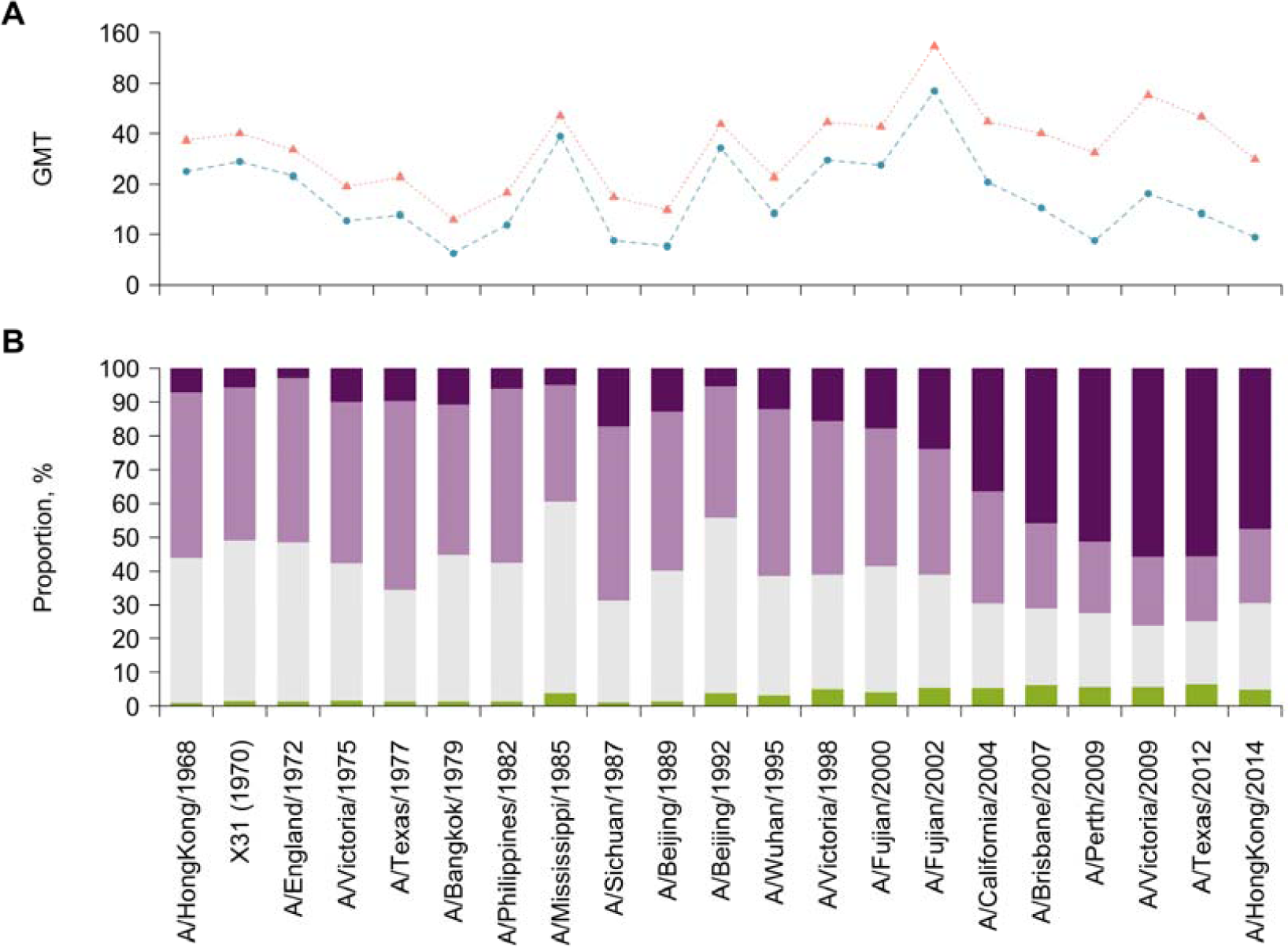
Changes in antibody profiles between visits. (**A**) Geometric mean titers (GMT) by strain. Blue and red points represent GMT at baseline and follow-up, respectively. (**B**) Distribution of changes in titers by strain. We divided the changes in titers into four categories, i.e. decrease (green), no change (grey), two-fold increase (light purple) and four-fold change (seroconversion, dark purple).

### Effect of pre-existing immunity on seroconversion to recent strains

Pre-existing, homologous titer to circulating strains is a well-described predictor of risk of seroconversion (*14*), which is confirmed in our study (Table 1 and tables S5 and S6). However, we hypothesized that including metrics of antibody profiles that integrate previous exposure history would further improve predictions of the risk of seroconversion. Therefore, we examined the association between the risk of seroconversion with age at baseline sampling, pre-existing, homologous titer to outcome strains (the *i*th strain), titer to the *i-1*th strain, and summary metrics (i.e. nAUC, nW_10_, nW_40_, and nATY) of strains up to and including the *i-2*th strain (“immunity of non-recent strains” hereafter). We found including nAUC or nW40 improved model fit (Table 1, figs. S6 and 7). After adjustment for pre-existing titer to strain *i*, we found pre-existing nAUC and nW_40_ were each positively associated with seroconversion (Table 1 and tables S3, S5 to S7). Taking A/Texas/2012 as an example, pre-existing titer to A/Texas/2012 was negatively associated with the risk of seroconversion (OR: 0.45; 95% CI, 0.35 to 0.57; Model 3 in Table 1), while there was an increased risk of seroconversion associated with higher pre-existing nAUC (OR: 1.21; 95% CI, 1.10 to 1.34) and nW40 (OR: 5.53; 95% CI, 2.15 to 14.57).

**Table 1.** Associations between pre-existing immunity and seroconversion to four recent strains.

|  | Adjusted odds ratio (95% confidence interval) |  |  |  |
| --- | --- | --- | --- | --- |
|  | A/Perth/2009 | A/Victoria/2009 | A/Texas/2012 | A/HongKong/2014 |
| **Model 1** |  |  |  |  |
| Age at sampling | 1.00 (0.99, 1.01) | 0.99 (0.98, 1.00) | 0.99 (0.98, 1.00)* | 1.00 (0.99, 1.01) |
| Titer to strain *i*<sup>a</sup> | 0.43 (0.34, 0.54)* | 0.51 (0.42, 0.60)* | 0.49 (0.38, 0.61)* | 0.70 (0.56, 0.85)* |
| Titer to strain *i-I*<sup>a</sup> | 1.30 (1.10, 1.55)* | 1.03 (0.87, 1.22) | 1.06 (0.85, 1.32) | 0.93 (0.79, 1.09) |
| Deviance explained | 7.5% | 12.9% | 12.6% | 4.0% |
| **Model 2** |  |  |  |  |
| Age at sampling | 1.01 (1.00, 1.02) | 1.00 (0.99, 1.01) | 1.00 (0.99, 1.01) | 1.00 (0.99, 1.01) |
| Titer to strain *i* | 0.42 (0.33, 0.53)* | 0.48 (0.40, 0.57)* | 0.45 (0.35, 0.57)* | 0.67 (0.54, 0.82)* |
| Titer to strain *i-I* | 1.21 (1.02, 1.44)* | 0.97 (0.82, 1.16) | 0.97 (0.78, 1.22) | 0.85 (0.71, 1.01) |
| AUC<sup>b</sup> | 1.14 (1.05, 1.24)* | 1.17 (1.07, 1.29)* | 1.21 (1.10, 1.34)* | 1.17 (1.07, 1.29)* |
| Deviance explained | 8.3% | 14.0% | 14.0% | 5.0% |
| **Model 3** |  |  |  |  |
| Age at sampling | 1.00 (0.99, 1.02) | 1.00 (0.99, 1.01) | 1.00 (0.99, 1.01) | 1.00 (0.99, 1.01) |
| Titer to strain *i* | 0.43 (0.34, 0.54)* | 0.49 (0.40, 0.58)* | 0.45 (0.35, 0.57)* | 0.68 (0.55, 0.84)* |
| Titer to strain *i-I* | 1.24 (1.04, 1.47)* | 0.98 (0.83, 1.17) | 1.00 (0.80, 1.25) | 0.88 (0.74, 1.05) |
| Width, cut off 1:40<sup>b</sup> | 2.86 (1.20, 6.92)* | 4.50 (1.77, 11.71)* | 5.53 (2.15, 14.57)* | 2.79 (1.13, 6.98)* |
| Deviance explained | 8.0% | 13.9% | 13.8% | 4.5% |

<sup>a</sup> Strain *i* refers to the strain that was examined for seroconversion, and strain *i-I* refers to the most recent strain isolated prior to strain *i*. E.g. when using seroconversion to A/Perth/2009 as outcome, strain *i* and *i-I* will be A/Perth/2009 and A/Brisbane/2007, respectively.
<sup>b</sup> Metrics were calculated using titers to strains isolated after the birth of the participants and before the year that strain *i-I* was isolated. Adjustment was then performed by standardizing the metrics with the number of post-birth strains.
\* Statistical significant level of 0.05.

### Life course exposures continually shape antibody profiles of influenza

We found an individual’s antibody profile is dynamically updated across time. Based on the impact of seroconversion and transient antibody dynamics, we expected that titers to recent strains would vary over the course of our study, but that, whereas titers to non-recent strains would to be relatively constant and when variation did exist, it would be in a random direction. However, we found that antibody titers to non-recent strains overwhelmingly increased (Figs. 2F and 4B), inconsistent with random variation (fig. S8C) suggesting that most increases, even those to non-recent strains, can be attributed to exposure to circulating strains. Though titer to non-recent strains in general increased between the two samples, the shape of each person’s antibody profile was highly consistent between visits (median Pearson’s correlation 0.90; IQR, 0.82 to 0.94). This result is similar to previously described results from 5 landscapes for 69 individuals (*7*), suggesting that individuals maintain patterns of relative titer across historical strains, even while acquiring antibodies due to new exposures.

We expect individuals of different ages to show different antibody profiles due to differences in lifetime influenza exposures. However, we found that our summary metrics did not monotonically increase with age. There is a gradual increase in the overall (nAUC) and breadth of immunity (nW_40_) throughout childhood consistent with the accumulation of exposures (Fig. 3, J and K and fig. S1) (*3*–*5*). The observed decrease in nAUC and nW_40_ among people aged 40 to 50 and age-independent changes in these metrics (Fig. 3 D and H) could suggest low HAI antibody among this age group, which could be due to high non-specific immune responses [e.g. antibodies against HA stalk and neuraminidase (NA) and cellular immune responses; (*18*–*22*)]. These non-specific immunities, which may accumulate over multiple exposures and are not measured in our assay, could preventing people from being infected and producing updated, strain specific HA responses (*19, 20, 23*).

Our hypotheses about the mechanisms driving age patterns in antibody profiles implicitly assume repeated, regular exposure to influenza throughout life. Consistent with this assumption, our findings during this four-year period suggested a nearly universal exposure to H3N2 in this cohort (Fig. 2C and Fig. 3D), agreeing to observations of high H3N2 incidence during this period with particularly large numbers of cases in 2011-12 (*24, 25*).

### Future immune responses to influenza are driven by antibody profiles

Our work further quantified the complex role that antibody profiles play in future immune responses to influenza. We confirmed the prominent role of pre-existing, homologous titers in determining the risk of seroconversion. However, we found that participants who had higher immunity to previously exposed strains were more likely to experience seroconversion to recent strains after adjusting for homologous titer. These findings were not affected by age or self-reported vaccination (figs. S9 and S10 and tables S7 to S9). Given individuals with high immunity to non-recent strains tended to have higher homologous titers (fig. S11 and table S10 and S11), we investigated whether the effect of immunity to non-recent strains was mediated by homologous titer (figs. S12 and S13; detailed in Materials and Methods) and found that pre-existing immunity to non-recent strains imposes a positive direct effect on the risk of seroconversion to recent strains but a negative, homologous titer mediated, indirect effect (figs. S12 and S13; detailed in Materials and Methods).

The mechanism behind the positive association between immunity to non-recent strains and seroconversion to recent strains is unclear. One plausible hypothesis is that a subset of people tend to have a more vigorous titer response across strains (e.g. individual heterogeneity in immune responses). This is supported by our data that positive association can still be detected when using titer to an older strain, instead of the summary metric of antibody profiles, as the proxy of immunity to non-recent strains (fig. S14). Another plausible hypothesis is that non-HAI immunity that was acquired from previous infections (e.g. antibody to HA stalk and NA and cellular immune responses) could blunt the production of strain-specific antibody upon exposure to circulating strains (*18*–*21*), thus reducing the HAI titers to the circulating strains and increasing the probability of seroconversion. High antibody to non-recent strains could indicate individuals who have not experienced infection in recent times [e.g. low strain-specific antibody to currently circulating strains due to temporary protection from last infection; (*23, 26, 27*)]. Homologous HAI titers may reflect the combined binding ability of antibody targeting the tested antigen as well as antibody targeting previously encountered epitopes, and high antibody titers to non-recent strains may indicate particular distributions of these two that are less protective.

### Life course immunity of influenza provides new opportunities in influenza studies

Previous work on mapping antigenic distance of evolving strains and characterization of life course immunity has changed how we approach studies of immunity to influenza (*2, 7*). The summary metrics we developed allow efficient characterization of profiles that are amenable to broad use in the analysis. With these metrics, we integrated information of life course HAI antibody mediated immunity to influenza that accounted for complex exposure histories and cross-reactions between influenza strains. These metrics should help to quantify the life course of non-HAI immunity that were not measured by our assay, and the interactions between infection events and immunity to influenza through life.

## Conclusion

We developed multiple summary metrics to integrate information of antibody profiles, which enabled us to further demonstrate clear age patterns of these profiles. We found accumulation of immunity to H3N2 during the first twenty years of life, followed by reductions among 40-50 years old. These age patterns in antibody profiles are likely to be driven by continual exposure to influenza; our study suggested nearly universal exposures during a four-year interval. Meanwhile, antibody profiles were found to provide more information than homologous titers in predicting temporal changes in influenza immunity. In particular, higher immunity to older strains was found associated with increased risk of seroconversion to the currently circulating strains, which mechanism remains unclear.

## Data Availability

All relevant data and code to reproduce the study findings will be available at (https://github.com/UF-IDD/Fluscape_Paired_Serology).

## Author contributions

Conceptualization: J.L., C.Q.J., J.M.R., K.O.K, Y.G., S.R.; Data curation: B.Y., J.L., R.S.; Formal Analysis: B.Y., J.L., D.A.T.C.; Funding acquisition: D.A.T.C, J.L., J.M.R., C.Q.J., Y.G. and S.R. Investigation: R.S., Z.H., Y.G.; Methodology: B.Y., J.L., D.A.T.C.; Supervision: D.A.T.C. Visualization: B.Y., J.L., D.A.T.C.; Writing - original draft: B.Y., J.L., D.A.T.C.; Writing - review, and editing: B.Y., J.L., H.Z., C.Q.J., J.M.R., J.A.H., K.O.K., R.S., Y.G., S.R., D.A.T.C..

## Data and material availability

All relevant data and code to reproduce the study findings are available at (https://github.com/UF-IDD/Fluscape_Paired_Serology).

## Ethics statements

Study protocols and instruments were approved by the following institutional review boards: Johns Hopkins Bloomberg School of Public Health, University of Florida, University of Liverpool, University of Hong Kong, Guangzhou No. 12 Hospital, and Shantou University. Written informed consent was obtained from all participants over 12 years old; verbal assent was obtained from participants 12 years old or younger. Written permission of a legally authorized representative was obtained for all participants under 18 years old.

## Supplementary Materials

Materials and Methods

Figures S1-S19

Tables S1-S13

References (*28-29*)

## Supplementary Materials for

## Materials and Methods

### Cohort profile

The Fluscape study is an ongoing cohort study in and around Guangzhou City in southern China that collects sera from consenting participants at regular intervals (roughly annually), as well as information on demographics. The methods and study population have been described in detail elsewhere (*17*). Briefly, 40 locations were randomly selected from a spatial transect extending to the northeast from the center of Guangzhou. In each location 20 houses were randomly selected and all consenting residents over 2 years of age were enrolled in the study. When households left the study, they were replaced by new randomly selected households to maintain a population of 20 households per location.

We collected serum from participants at two time points, i.e. baseline (2009 to 2011) and follow-up (2014-2015). Serum from both visits were available from 777 participants out of 2,767 total participants (across both visits, table S1), of which 763 participants had interpretable titers for all 21 H3N2 strains (Fig. 2A-C, 14 participants had uninterpretable results for 5 strains). Age at the time of the baseline ranged from 2 to 86 years old, with over representation of participants aged 40-59 years compared to the Chinese population according to the 2009 census (51.4% vs. 31.6%) (*17*).

### Laboratory testing

Blood samples were kept at 4°C until processing on the day of collection. Serum is extracted from these samples and frozen at -80°C until testing. For all individuals recruited at both baseline visit (4 December 2009 to 22 January 2011) and follow-up visit (17 June 2014 to 2 June 2015) we measured hemagglutination inhibition (HAI) titers for antibodies against a panel of 21 strains of H3N2 influenza spanning the history of the virus since its emergence in humans in 1968 (A/Hong Kong/1968, X31, A/England/1972, A/Victoria/1975, A/Texas/1977, A/Bangkok/1979, A/Philippines/1982, A/Mississippi/1985,A/Sichuan/1987, A/Beijing/1989, A/Beijing/1992, A/Wuhan/1995,A/Victoria/1998, A/Fujian/2000, A/Fujian/2002, A/California/2004, A/Brisbane/2007, A/Perth/2009, A/Victoria/2009, A/Texas/2012, A/Hong Kong/2014). For each strain-individual pair, HAI titers were determined by two-fold serial dilutions from 1:10 to 1:1280 conducted in 96-well microtiter plates with 0.5% turkey erythrocytes. Sera from the two visits were tested side by side on the same plate, and confirmatory samples collected in 2014 sera were tested on a separate plate. The reciprocal of the highest dilution where hemagglutination does not occur is reported as the titer. There were 28 missing (for 14 individuals and 5 strains) out of 32,636 strain-individual-visit combinations, which were due to inadequate sera remaining or inconclusive readings.

### Analytic methods

For analysis, individuals with undetected titers are assumed to have a titer of 5, and all titers are transformed to the log-scale. Specifically, we transform titers based on the formula y=log2(x/5) where x is the measured titer, which results in a 0 for undetectable titers and a 1 unit rise in titer for each 2-fold increase (so 10=1, 20=2, 40=3, etc.). In figures, measures are transformed back to the arithmetic scale for clarity; but all statistics are presented on this scale. Geometric mean titer (GMT) was calculated as the mean of log-titer and transformed back to the arithmetic scale. 95% confidence intervals (CIs) were calculated using student t tests assuming the log-titer follows a normal distribution. Age at baseline sampling is defined as the time difference between the year of baseline and the year of birth; age at isolation is calculated as the year when the tested strain was isolated minus the year of birth of the participant.

#### Summary metrics

Individual antibody profiles were constructed using each individual’s HAI titers against 21 tested H3N2 strains which were sorted in chronological order. We characterized the shape of these individual antibody profiles using a number of statistics (Fig. 3) which are proposed to approximately estimate the area under the antigenic landscape surface (Area Under the Curve, AUC), the breadth of antibody profile (Width, W), where the breadth is calculated above either detective (1:10) or protective threshold (1:40), and the temporal targeting of antibody profile by the measured HAI titer values (Averaged Titer Year, ATY). The statistics are calculated as follows:

1. *Area Under the Curve (*AUC): We estimated the area under the curve of antibody profiles as:

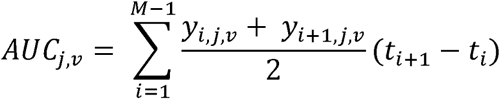 Where *AUC*_*j,V*_ is the area-under the curve of titers by time for person *j* and visit *v, M* is the total number of included strains, *y*_*i,j,V*_ is participant *j*‘s log-titer against strain at visit *v*, and *t*_*i*_ is the time of isolation of strain *i*.
2. *Width (*W): We define the width of the curve to be the proportion of time during which the antibody profile that is greater than or equal to some predefined antibody titer cutoff, z. Here we focus on detectable titers (*W*_10_, *z* = 1:10), and protective titers (*W*_40_), based on the commonly used cutoff. Hence,

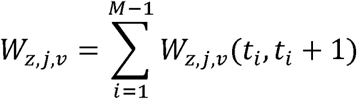

where:

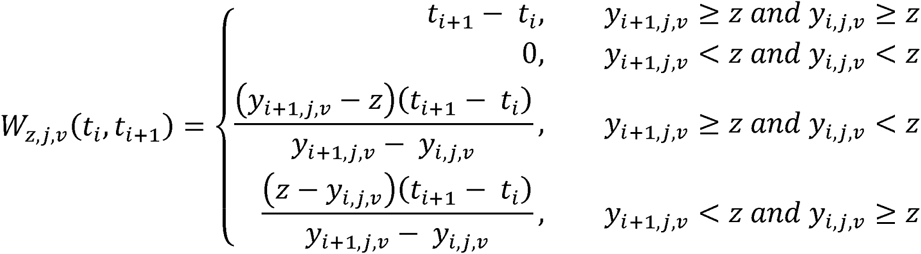 When titers to all strains are above the threshold *z*, the width for an individual given the tested strains is at its maximum:

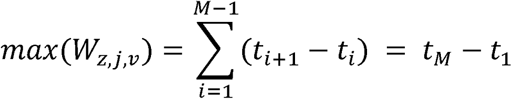 We present the width (ranges from 0 to 1) standardized by the maximum width in the results:

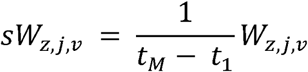
3. *Average Titer Year (A*TY): The average titer year is the center of mass of the curve with respect to strain isolation time, capturing all of an individual’s titer values (i.e., the weighted average of their titers):

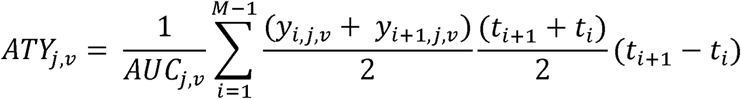

In order to account for the impact of pre-birth strains, we calculated the metrics using post-birth strains and normalized with the number of post-birth strains (*M*_*post*_) in the main analysis. Exclusion of pre-birth strains was performed for each participant according to the year of birth. The normalized statistics are calculated as follows:

1. *Normalized Area Under the Curve (nAUC)*:

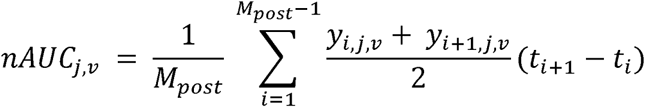
2. *Normalized Width (nW)*:

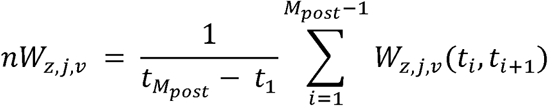
3. *Normalized Average Titer Year (nATY)*:

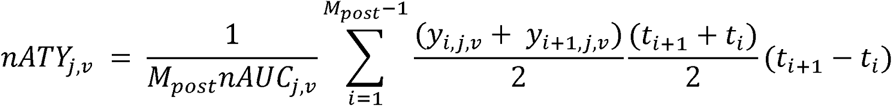

For both normalized and unnormalized metrics, the changes in area under the curve and width between the visits were calculated as the difference between baseline and follow-up values of these metrics. In order to examine the temporal targeting of the changes in titers between the two visits, the changes in nATY (Fig. 3L) and ATY (fig. S2L) are calculated using difference in titers:

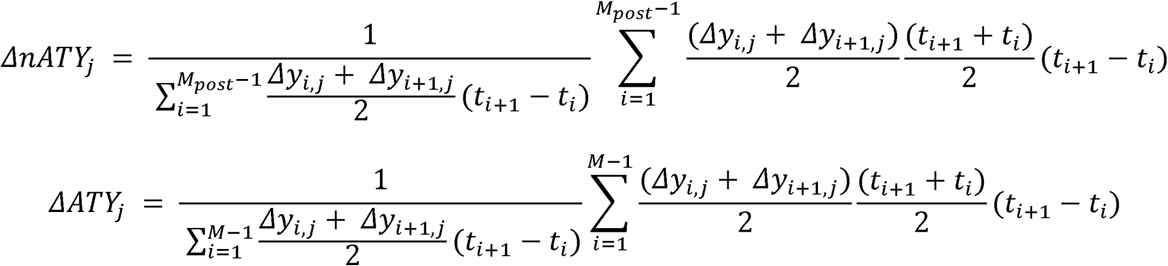

#### Age pattern in antibody profiles

To account for the heterogeneous exposure history of participants at different ages (Fig. 2G-I), we fitted generalized additive models (GAM) to the measured HAI titers against all tested H3N2 strains incorporating a smoothed interaction term of age at sampling and year of the strains circulated. We also fitted separate GAM to examine the non-linear association between the each of summary analytic statistics mentioned in the above section and the ages of participants at baseline sampling (Fig. 2 and figs. S2 to S3).

#### Changes in antibody profiles between visits

The change of HAI titer is defined as the difference in the measured log-titers against tested H3N2 strain between the baseline and follow-up visits. We characterized the strain distribution among titers that were decrease, no change, two-fold increase and four-fold increased (seroconversion), respectively and compared such distribution with the underlying strain distribution (i.e. the number of titers of that strain divided the number of titers of all tested strains) (fig. S5). We fitted logistic regression of seroconversion (*c*_*i,j*_), on strains (*s*_*i*_ *as a categorical variable*) and adjusted for age at baseline sampling (*a*_*j*,1_) and prior log-titers (*y*_*i,j*,1_), to further examine the strain-specific risk of seroconversion (*α*_3_, fig. S4):

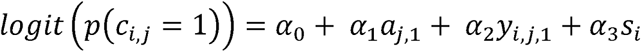

We characterized the changes of HAI titers to four recent strains that were possibly circulating during our study period, i.e. A/HongKong/2014, A/Texas/2012, A/Victoria/2009 and A/Perth/2009. Distribution of changes by the number of strains that participants showed increased titers to and by individual strain was characterized, respectively (fig. S5). 95% confidence intervals were calculated by assuming a binomial distribution.

#### Effects of pre-existing immunity on seroconversion to recent strains

We predicted the risk of seroconversion (*c*_*i,j*_) to one of four recent strains *i* (i.e. A/HongKong/2014, A/Texas/2012, A/Victoria/2009 or A/Perth/2009) by fitting logistic regression with predictors that reflect varying durations of exposure history and adjusting for age at baseline sampling (*a*_*j*,1_). In brief, we progressively included log-titer to the examined strain (*i*th, *y*_*i,j*,1_), log-titer to the strain prior to the examined strain (*i-1*th, *y*_i−,*j*,1_) and one of the summary metrics (*I* _*j*,1_ ∈{*AUC*_*j*,1_, *ATY*_*j*,1_, *W*_*z,j*,1_}) that was calculated using strains isolated since the year of birth (or 1968 when including pre-birth strains) to the year before the *i-1*th strain was isolated. The model adjusting for the most complete exposure history is:

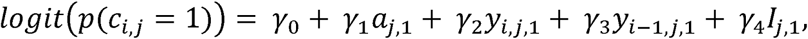

and the model was fitted to each recent strain separately. Akaike information criterion (AIC) and Bayesian information criterion (BIC) were used to compare the performance of the models (figs. S6 and S7).

All analyses were performed using R version 3.5.0 (R Foundation for Statistical Computing, Vienna, Austria), among which we fitted GAM using “mgcv” package (*27*).

### Additional analysis

#### Age patterns in strain-specific titers

We fitted univariable GAM to the measured log-titers on the participants’ age at baseline sampling (i.e. participant age when serum was collected at baseline sampling) and age at isolation [i.e. participants’ ages when strains were isolated; (*4*)], respectively to examine the strain-independent age patterns of HAI titers (fig. S1). The relationship between age at sampling and HAI responses varied across strains (fig. S1 A and B). However, we found consistent patterns in the relationship between participant age at isolation and HAI responses, particularly the age at isolation for strains against which people have the highest titers (fig. S1 D and E).

Median participant age at the time of isolation of strains to which they had the highest HAI titers was 4.3 years (IQR: 2.0 to 6.9 years across strains). As age at isolation increases, titers decrease rapidly until 20 years of age.

#### Features of non-normalized summary metrics of antibody profiles

Results of AUC that included pre-birth strains (i.e. isolated before participants were born) showed similar age patterns to including only post-birth strains, except a lower ratio of average AUC among participants aged ≤20 years old to that among 40-70s (0.96 for baseline and 0.82 for follow-up) (fig. S2 A and B). This is due to the relatively shorter exposure history of participants aged ≤20 years old compared to that among 40-70s (i.e. a smaller number of strains that could have been exposed to), and the lower titers to pre-birth strains (Fig. 2 A and B and fig. S1). We also divided the H3N2 strains into four subgroups following a chronological order and calculated AUC of strains in each subgroup. Similar to findings in fig. S1, AUC peaked among participants who were 10 years old when strains were isolated (fig. S16).

Non-normalized W_40_ illustrates an increasing trend as participants’ age grows until ∼30 years old, which is older than the peaking age of nW_40_ (fig. S2 D and E). Shorter exposure history of young children and lower responses to pre-birth strains contributed to the lower W_40_ compared to nW_40_. As a result, the drop in W_40_ among 40 - 70 years old is less compared to that in nW_40_.

Results of ATY that included pre-birth strains showed almost identical pattern to that of nATY (fig. S2 J, K and L).

#### Ceiling effects of strain distribution by changes in titers

We characterized the strain distribution among for subgroups of titers, i.e. decrease, no change, two-fold increase and seroconversion and compared these distributions with the underlying strain distribution (fig. S15). The underlying strain distribution was calculated by dividing the number of available titers for the strain by the total number of available titers for all strains in the examined dataset. In order to assess the ceiling effects, we performed this analysis to two separate subsets of titers that only include pre-existing titers ≤ 1:80 and > 1:80 (fig. S15 B and C), respectively. Results suggested that the strain distribution by changes in titers seemed not to be affected by the ceiling effects, with the most dynamic changes observed for recent strains.

#### Age distributions by changes in titers

We characterized the age distribution among titers that showed varying changes, i.e. decrease, no change, two-fold increase and four-fold increase (fig. S8). We compared these distributions with the underlying age distribution of participants (table S1) who provided sera using Fishers’ exact test. There appeared to be a higher proportion of people aged 10-20 years old (14.4%; 95%CI, 9.0% to 21.3%) and a lower proportion of people aged 40-50 years (23.0%; 95% CI, 16.3% to 30.9%) of participants who experienced decreased titers compared to all participants (Fisher’s exact test, P = 0.03, fig. S8E). There was no difference in the age distributions of participants who had unchanged, any-fold increased or seroconversion and the underlying age distribution of participants (Fisher’s exact test, P > 0.05, fig. S8 F and H).

#### Effects of pre-existing titers on changes in titers

To examine the effect of pre-existing titers on the changes in HAI titers, we fitted linear regression of changes in log-titers (*Δ,y*_*i,j*_) on the pre-existing log-titers to single strain *i* for participant *j* strains (*y*_*i,j*,1_) and adjusted for age at baseline sampling (*a*_*j*,1_), tested H3N2 strains (*s*_*i*_) and the interaction term of H3N2 strains and pre-existing titers (*s*_*i*_ × *y*_*i,j*,1_):

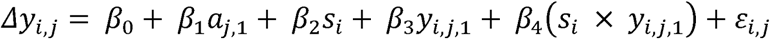

We assessed the strain-specific effects of pre-existing titer through the estimated coefficients for the strain and pre-existing titers interaction term (i.e. *β*_4_) (fig. S17, tables S12 and S13).

Pre-existing titers (*ΔR*^2^9.3%) and strains (*ΔR*^2^13.9%) explained more variations in changes of titers than age (*ΔR*^2^0.3%) (table S13). Overall, we found protective associations of pre-existing HAI titers on the changes of titers between baseline and follow-up and such associations were larger for more recently isolated H3N2 strains (fig. S17, tables S12 and S13).

#### Effects of pre-existing immunity on seroconversion to recent strains

##### Effects of nW10 and nATY on seroconversion to recent strains

Except for nW10 for A/Victoria/2009, we found positive associations between nW10 or nATY and seroconversion to recent strains after adjusting for age at sampling, titer to strain i and strain i-1 (table s4). However, including nW10 or nATY seems not always improve the model fit, especially when accounting for the non-linear effects of age (figs. S6 and S7).

##### Effects of non-normalized pre-existing immunity of non-recent strains on seroconversion to recent strains

When fitted logistic regressions of seroconversion on recent strains with age at sampling, titer to strain *i*, titer to strain *i-1* and non-normalized summary metric of antibody profiles, positive associations were only found for ATY for all four strains and W_40_ for A/Perth/2009, A/Victoria/2009 and A/Texas/2012 (tables S3 and S4). Non-normalized ATY and W40 were less likely to be affected by the pre-birth strains, as individuals are more likely to have weak but broad cross-reactions to strains that they could not have been exposed to.

##### Univariable analysis of effects of pre-existing immunity on seroconversion to recent strains

We performed univariable logistic regressions of seroconversion to each of the four recent strains on the covariates included in the main analysis, i.e. age at baseline sampling, titer to strain *i*, titer to strain *i-1* and each summary metric of antibody profiles. Only titers to strain *i* and strain *i-1* were found to have negative associations with seroconversion to all four recent strains (table S11). In addition, we found negative associations between seroconversion to a given strain that were isolated after 1992 and titers to its antigenically relatives (fig. S11A) from univariable logistic regressions of seroconversion of strain *a* on titer to strain *b*. However, such associations were not significant after accounting for the titer to the examined outcome strain (fig. S11B).

These results confirmed the protective effects of per-existing, homologous titer on seroconversion. We also found strong correlation between covariates that we included in the models to examine the effects of pre-existing immunity on seroconversion to four recent strains (table S10). We found titer to strain *i*, titer to strain *i-1* and each summary metric of antibody profiles were negatively associated with age at baseline sampling. The summary metrics of antibody profiles were positively associated with titer to strain *i*, which is likely driven by the cross-reactions.

##### Mediation analysis of the association between immunity of non-recent strains and seroconversion to recent strains

We did not find a positive association between pre-existing immunity to non-recent strains (i.e. nAUC or nW_40_) and seroconversion to recent strain when titer to strain *i* and titer to strain *i-1* (which is highly correlated with titer to strain *i*) were not adjusted (Model 3 in table S6), which is probably due to the positive correlation between pre-existing immunity to non-recent strains and titer to strain *i* (table S10). Therefore, we hypothesize that pre-existing immunity to non-recent strains imposes both indirect effects mediated by titer to strain *i* and a direct effect on seroconversion to strain *i*. In order to further separate the direct effect of immunity of non-recent strains from the indirect effect, we conducted a mediation analysis based on the causal diagram shown in fig. S12. We first fitted the mediation model using a linear regression of pre-existing log-titer to strain *i* (*y*_*i,j*,1_) on pre-existing immunity to previous exposures (*I*_*j*,1_) and adjusting for age at baseline (*a*_*j*,1_). We then fitted the outcome model using a logistic regression of seroconversion to strain *i* (*c*_*i,j*_) on pre-existing log-titer to strain *i* (*y*_*i,j*,1_), pre-existing immunity to previous exposures (*I*_*j*,1_) and age at baseline (*a*_*j*,1_). We estimated total effect, average direct effect and average indirect effect using the “mediation” package with 1,000 bootstrap samples (*28*). We also fitted the outcome model with an additional interaction term between pre-existing log-titer to strain *i* and pre-existing immunity to previous exposures in order to account for the effect of their interactions on the mediation effects (fig. 13).

Results suggested both nAUC and nW_40_ had negative indirect effects and positive direct effects (except for nW_40_ on A/HongKong/2014) on seroconversion to strain *i*, which yielded non-significant or marginally negative total effects (fig. S13). Results of mediation analyses were not affected by the possible interactions between pre-existing log-titer to strain *i* and pre-existing immunity to previous exposures (fig. S13).

##### Effects of age on the association between immunity of non-recent strains and seroconversion to recent strains

We assumed a linear effect of age on seroconversion to recent strains in the main analysis, and performed sensitivity analysis by assuming a non-linear effect of age. We fitted GAM with seroconversion to recent strains adjusted for age in a spline form, titer to strain *i*, titer to strain *i-1* and the summary metrics. We found consistent patterns in the association between age and seroconversion after adjusting for other covariates, however, these associations were not significant (fig. S9) and did not affect our main outcome (table S7).

##### Performance of predictions in seroconversion to recent strains

We compared the predicted probability of seroconversion to four recent strains with the observed proportion of seroconversion among age groups binned by 10 years, in order to examine the model performance in predictions. We examined the uncertainty of prediction with the interquartile of predicted probability of each age group and derived 95% CI of observed proportion of seroconversion assuming a binomial distribution. Predictions and observations showed good consistency, and both were with great uncertainty (figs. S18 and S19). There were two age groups with high predicted probability but low observed proportion of seroconversion when assuming a linear association between age and seroconversion (fig. S18), which was due to the edge effect of age that can be accounted by a spline term on age (figs. S9 and S19).

#### Effects of vaccination against influenza

We included serum from all participants regardless of their vaccination status in the main analysis and used serum from participants who self-reported no previous vaccination against influenza in both visits in the sensitivity analysis (table S18), in particular summary metrics metrics of antibody profiles and effects of pre-existing immunity on seroconversion to recent strains. Age patterns in summary metrics of antibody profiles of those unvaccinated participants (fig. S10) were similar to results including all participants (Fig. 2 and fig. S3). Vaccination status of influenza seems not affect the associations between pre-existing immunity and risk of seroconversion to recent strains (table S9).

## Supplementary Figures

**Fig. S1.**
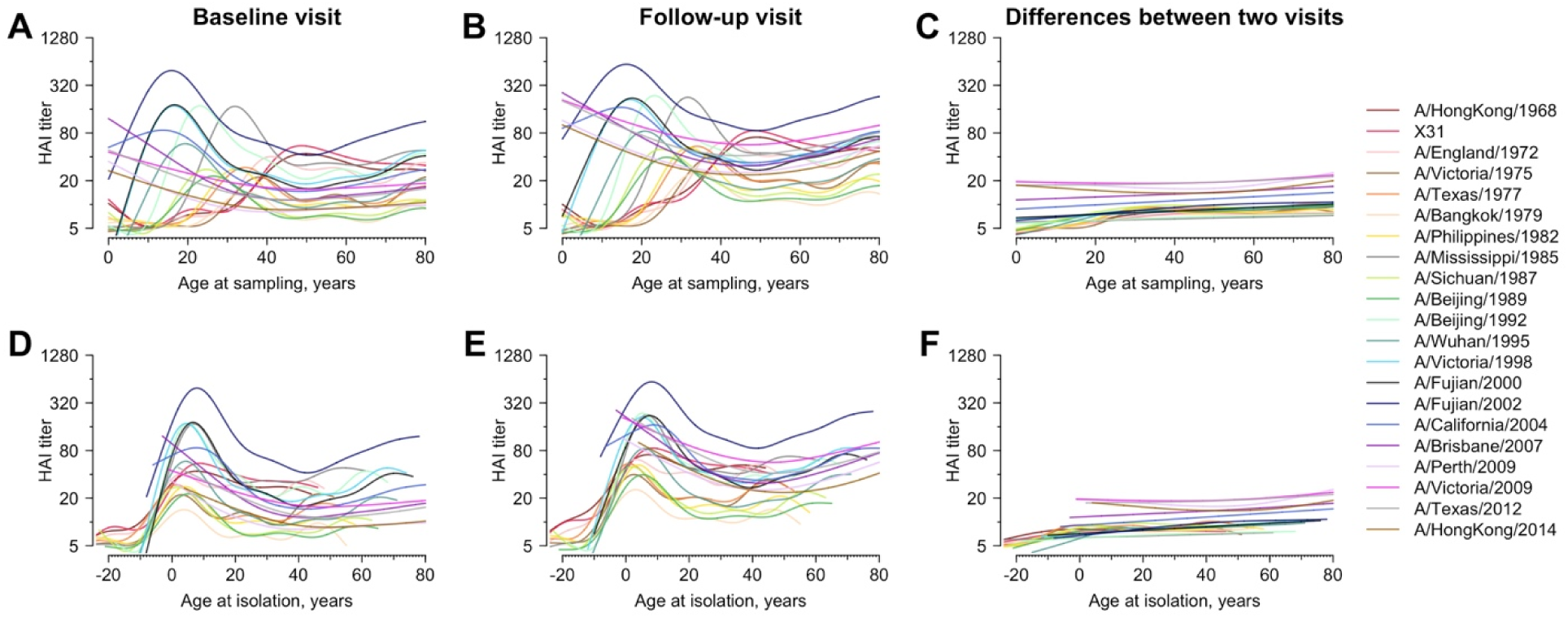
Age and HAI titer against individual H3N2 strain. Lines are the predicted mean HAI titer fitted from general additive model (GAM) using age at sampling (panel A to C) and age at isolation (panel D to F) as predictor, respectively. We fitted separate GAMs for HAI titers measured for serum collected from baseline (panel A and D), serum collected in follow-up visit (panel B and E) and the differences between two visits (panel C and F).

**Fig. S2.**
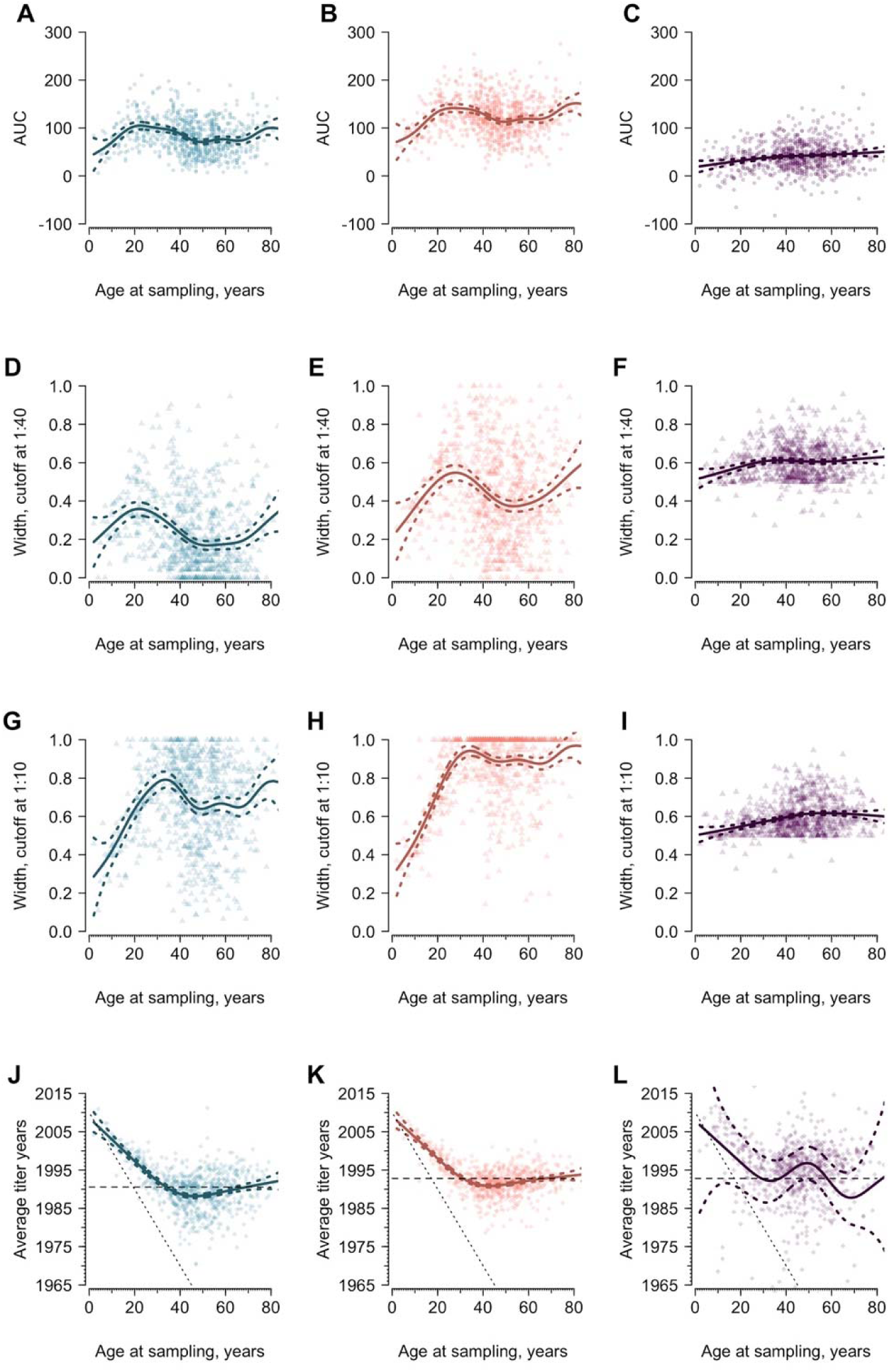
Area under the curve (AUC), average titer years (ATY) and width varying with age, using all tested strains. Blue and red represent the AUC for the baseline and follow-up visit, respectively. Purple indicates the differences of indicators between the two visits. Solid lines are predictions from gam and the colored dashed lines represent the corresponding 95% confidence intervals. The sloping black dotted lines in panel **J** to **L** indicate the year of birth of participants. The dashed lines in panel **J** to **L** indicate the unweighted average isolation year of post-birth strains.

**Fig. S3.**
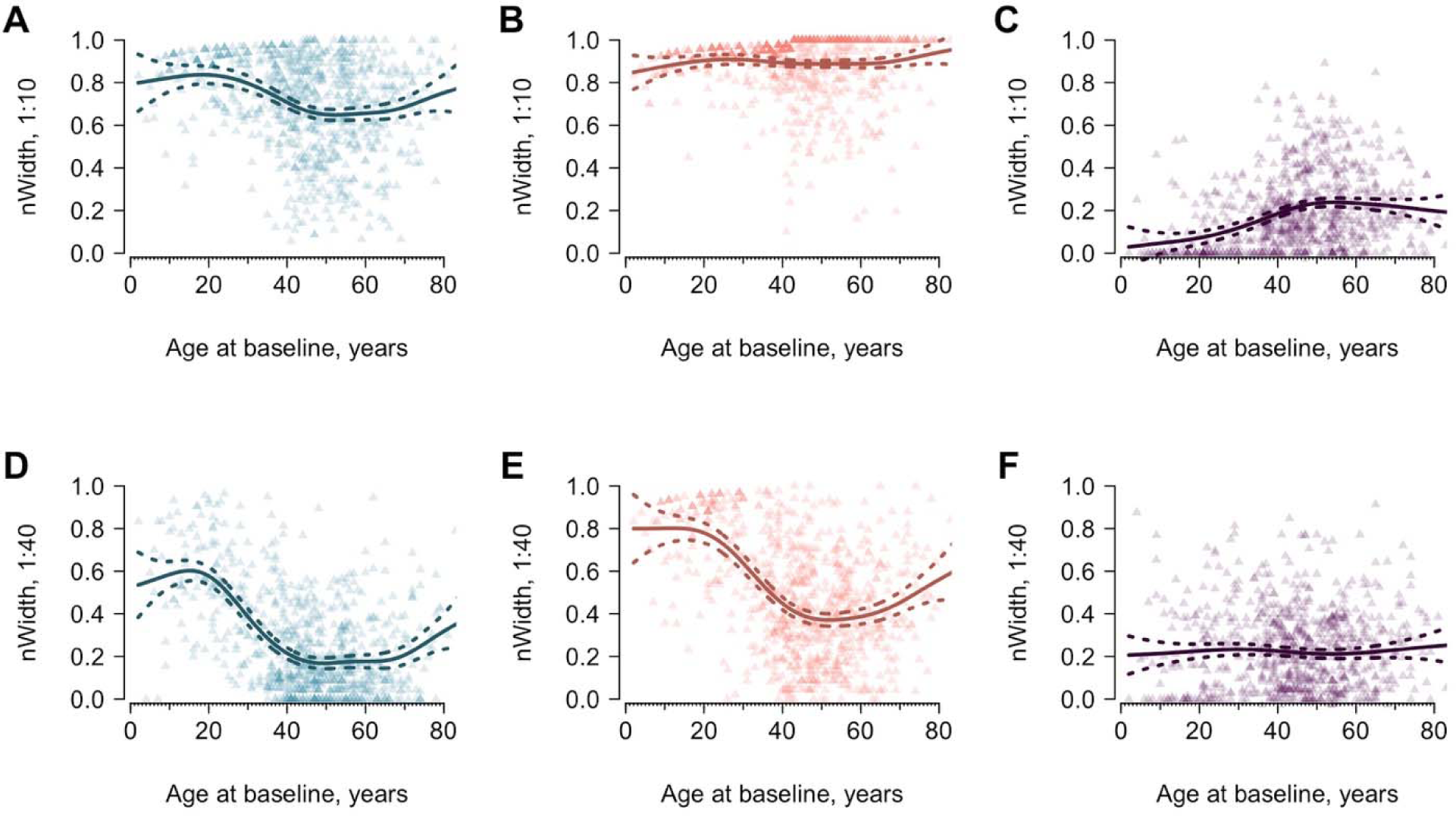
Width of antibody profiles varying with age. Widths were calculated using post-birth strains only. Panel **A** to **C** demonstrate width above titer 1:10, and Panel **D** to **F** demonstrate width above titer 1:40. Blue and red represent the indicators measured for serum collected in 2010 and 2014, respectively. Purple indicates the differences of indicators between the two visits. Solid lines are predictions from generalized additive model and the colored dashed lines represent the corresponding 95% confidence intervals. Results were calculated including all strains.

**Fig. S4.**
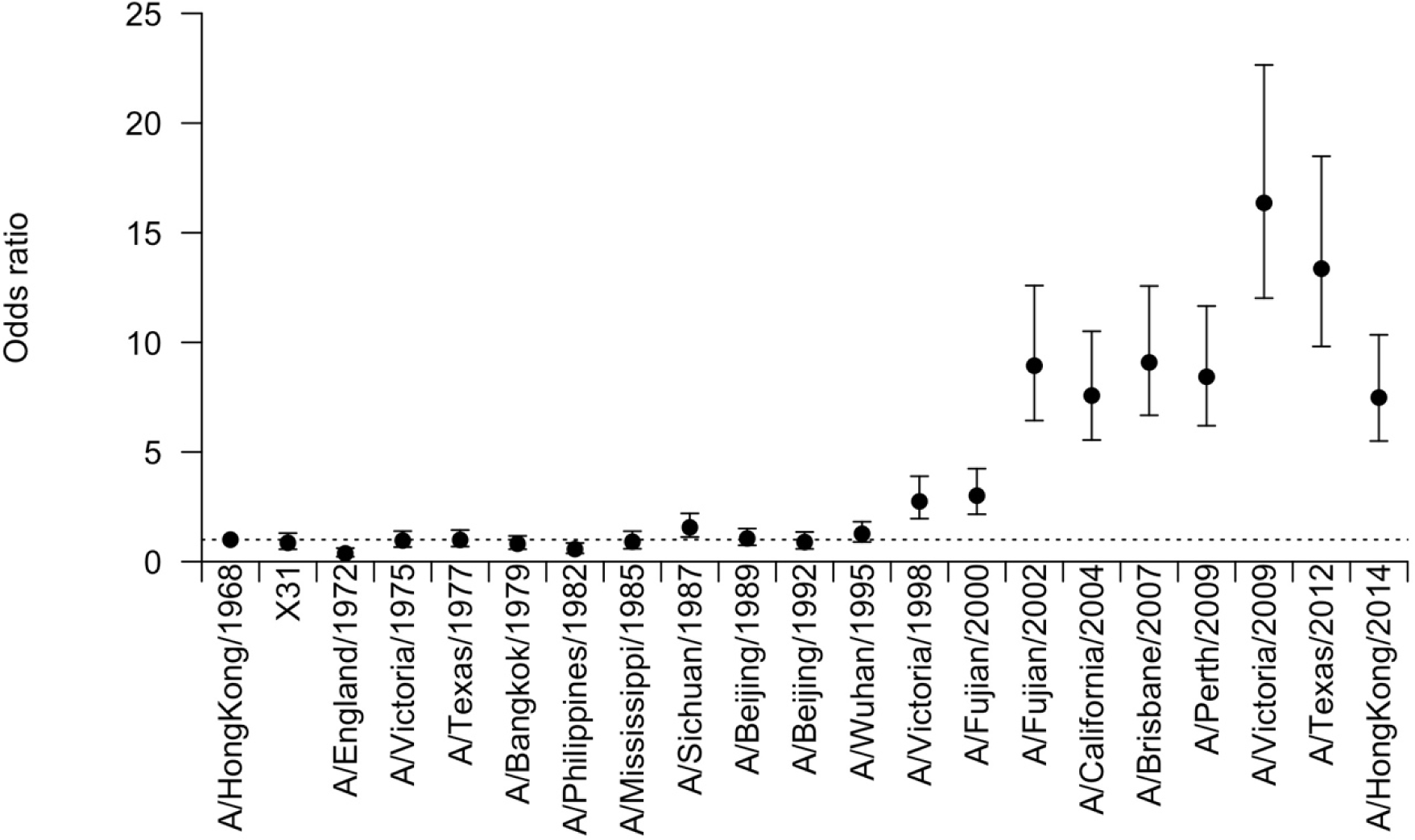
Risk of seroconversion by H3N2 strains. Logistic regression models were fitted using age at sampling, prior titer and strains to predict the seroconversion. Coefficients for H3N2 strains are shown in the figure.

**Fig. S5.**
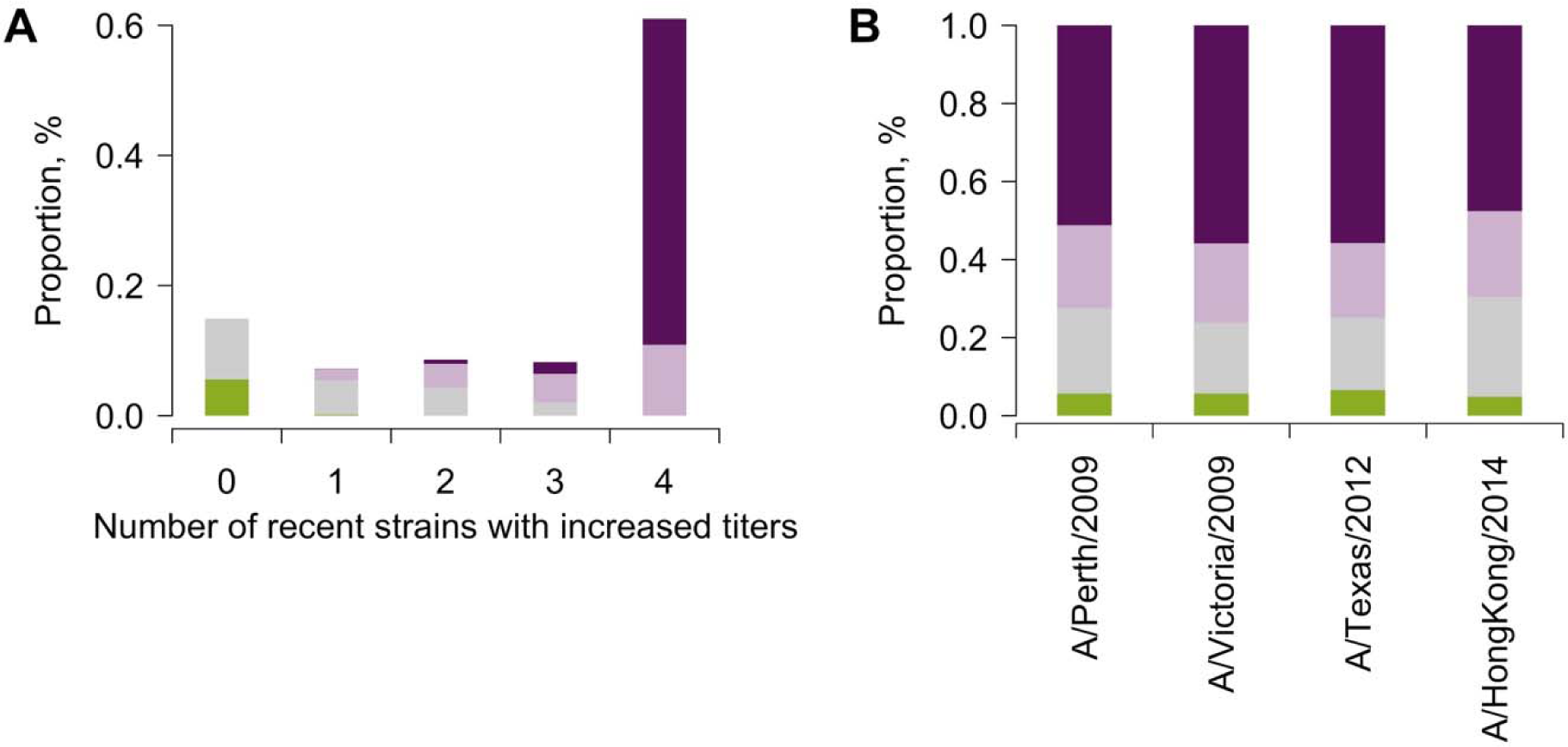
Changes in titers to four recent strains. (**A**) Distribution of changes in titers against recent H3N2 strains by the number of strains with increased titers. (**B**) Distribution of changes in tiers against recent H3N2 strains by individual strain.

**Fig. S6.**
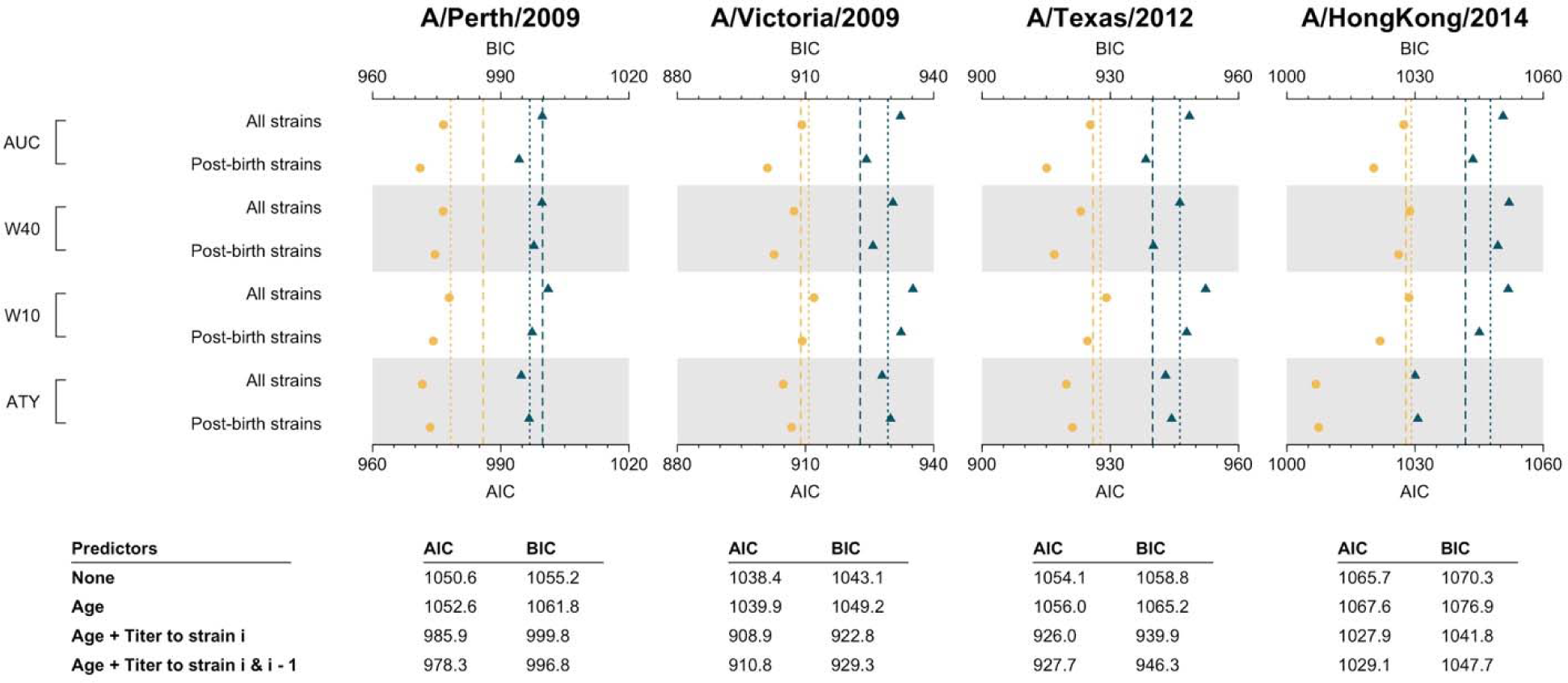
Comparison of prediction performance of models including pre-existing immunity. Yellow and blue represents AIC and BIC, respectively. Dashed lines represent the AIC/BIC for models that only included titer to the examined strain *i*. Dotted lines represent the AIC/BIC for models that included titers to the examined strain *i* and the prior strain *i-1*. Dots are AIC/BIC for models including additional predictor of pre-existing immunity of strains up to strain *i-1*.

**Fig. S7.**
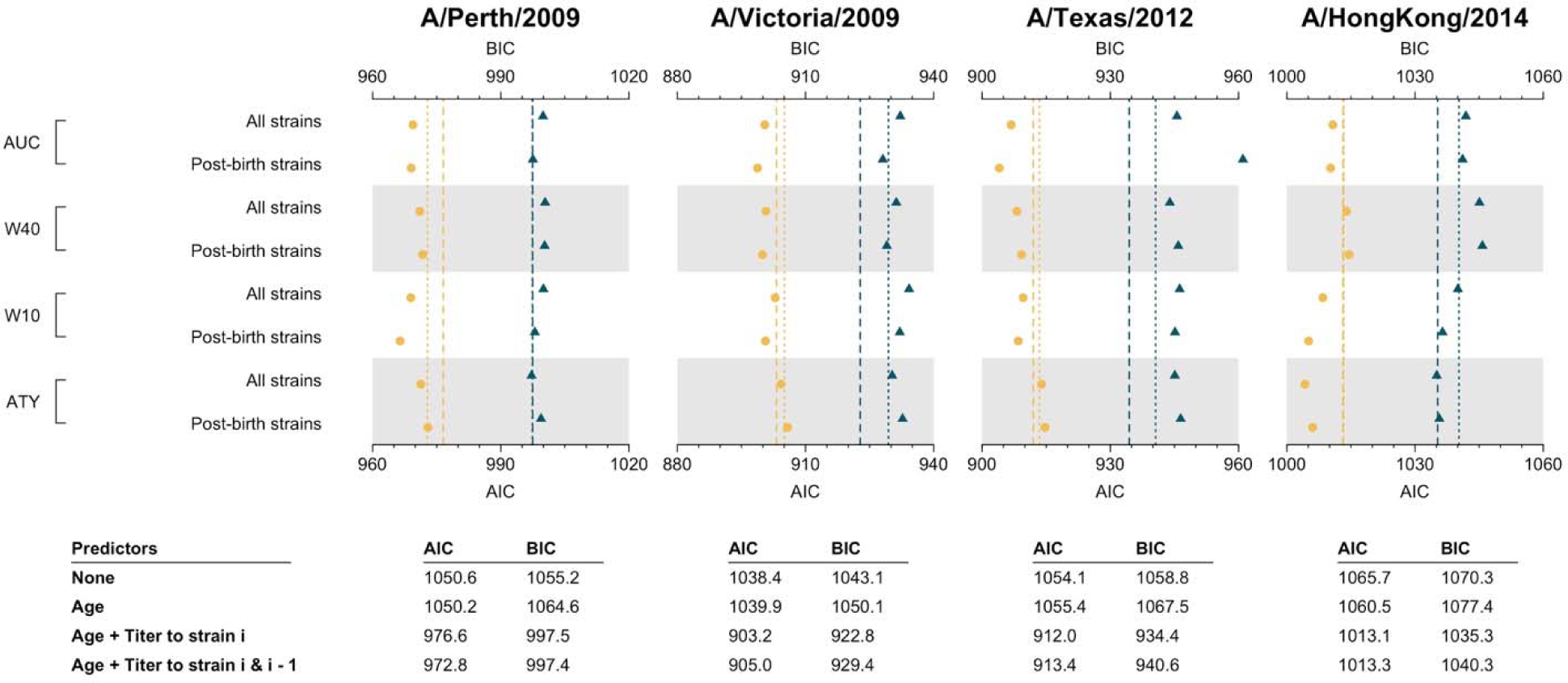
Comparison of prediction performance of models including pre-existing immunity. Yellow and blue represents AIC and BIC, respectively. Dashed lines represent the AIC/BIC for models that only included titer to the examined strain *i*. Dotted lines represent the AIC/BIC for models that included titers to the examined strain *i* and the prior strain *i-1*. Dots are AIC/BIC for models including additional predictor of pre-existing immunity of strains up to strain *i-1*.

**Fig. S8.**
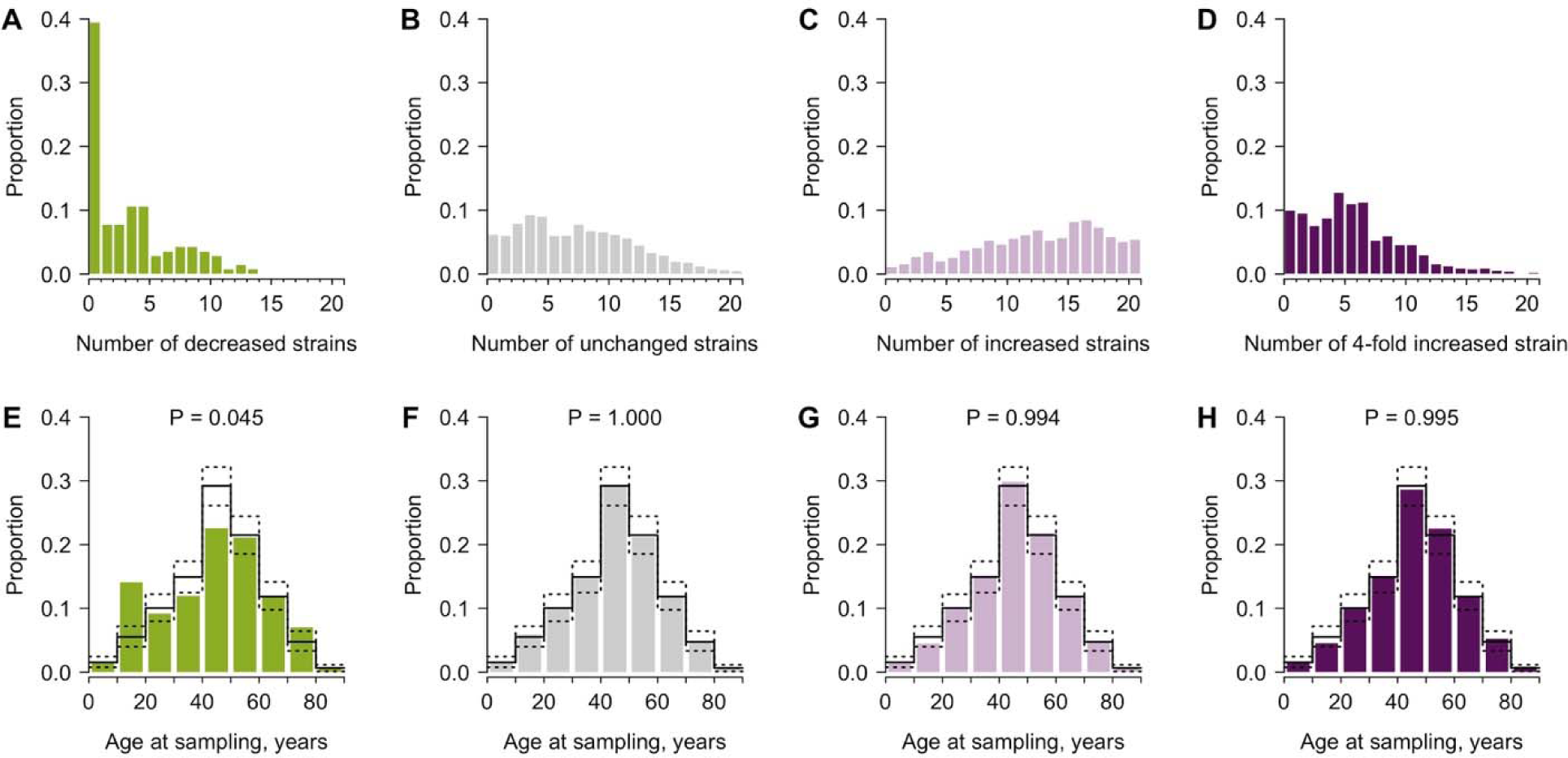
Age and differences of HAI titers between two visits. Columns from left to right shows results of subgroups of participants who had decreased, unchanged, increased and four-fold increased titers between the two visits, respectively. **E** - **H**: Solid lines were the age (at sampling) distribution of all participants who gave serum and dotted lines were the 95% confidence intervals derived from 1,000 bootstraps. P values shown were derived from Fisher’s exact test for the age distribution of the corresponding subgroup and the age distribution of all 777 participants.

**Fig. S9.**
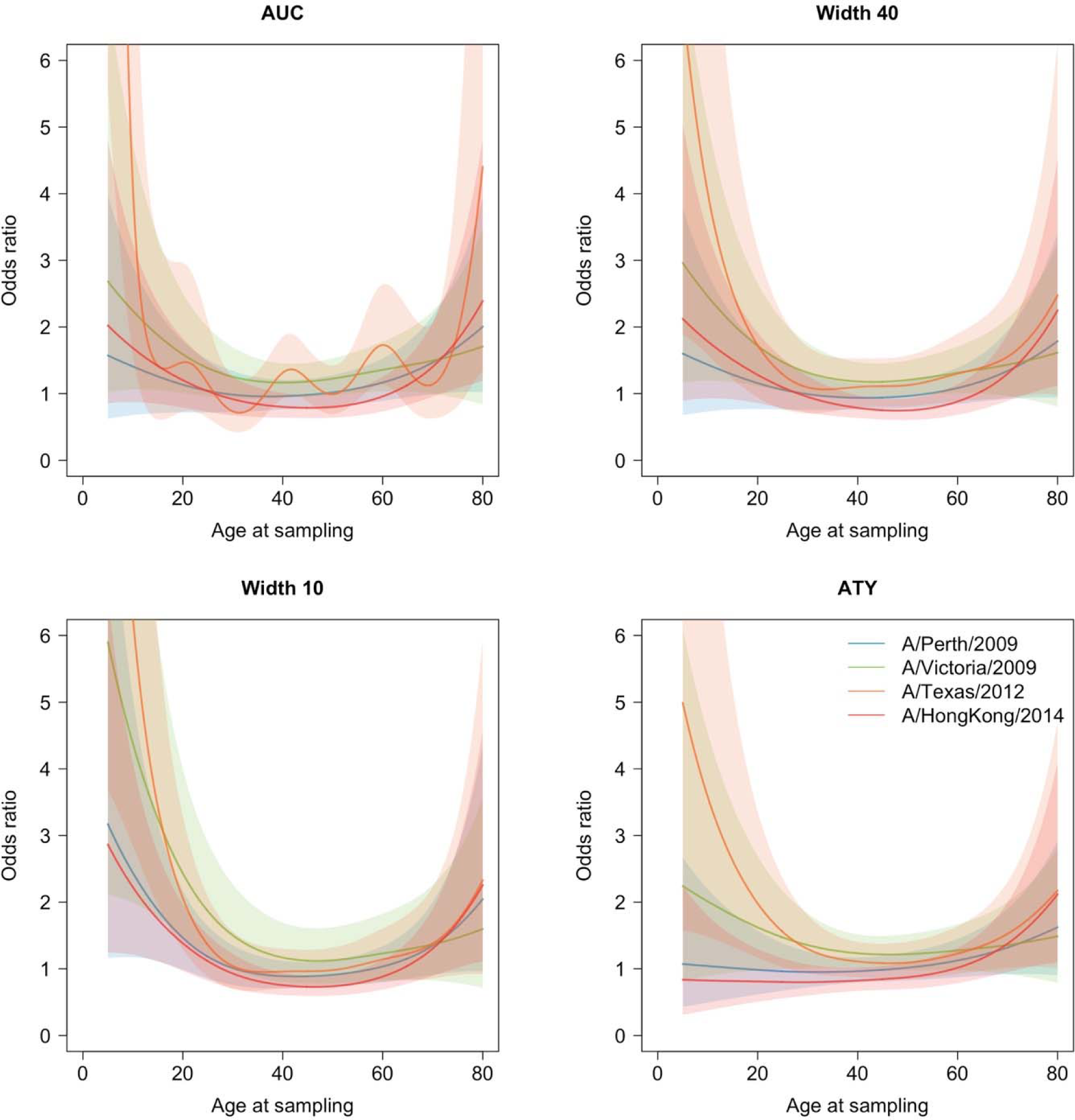
Non-linear associations between age at sampling and seroconversion of four recent strains. Models has been adjusted for titer to strain *i*, titer to strain *i-1*, and summary metrics.

**Fig. S10.**
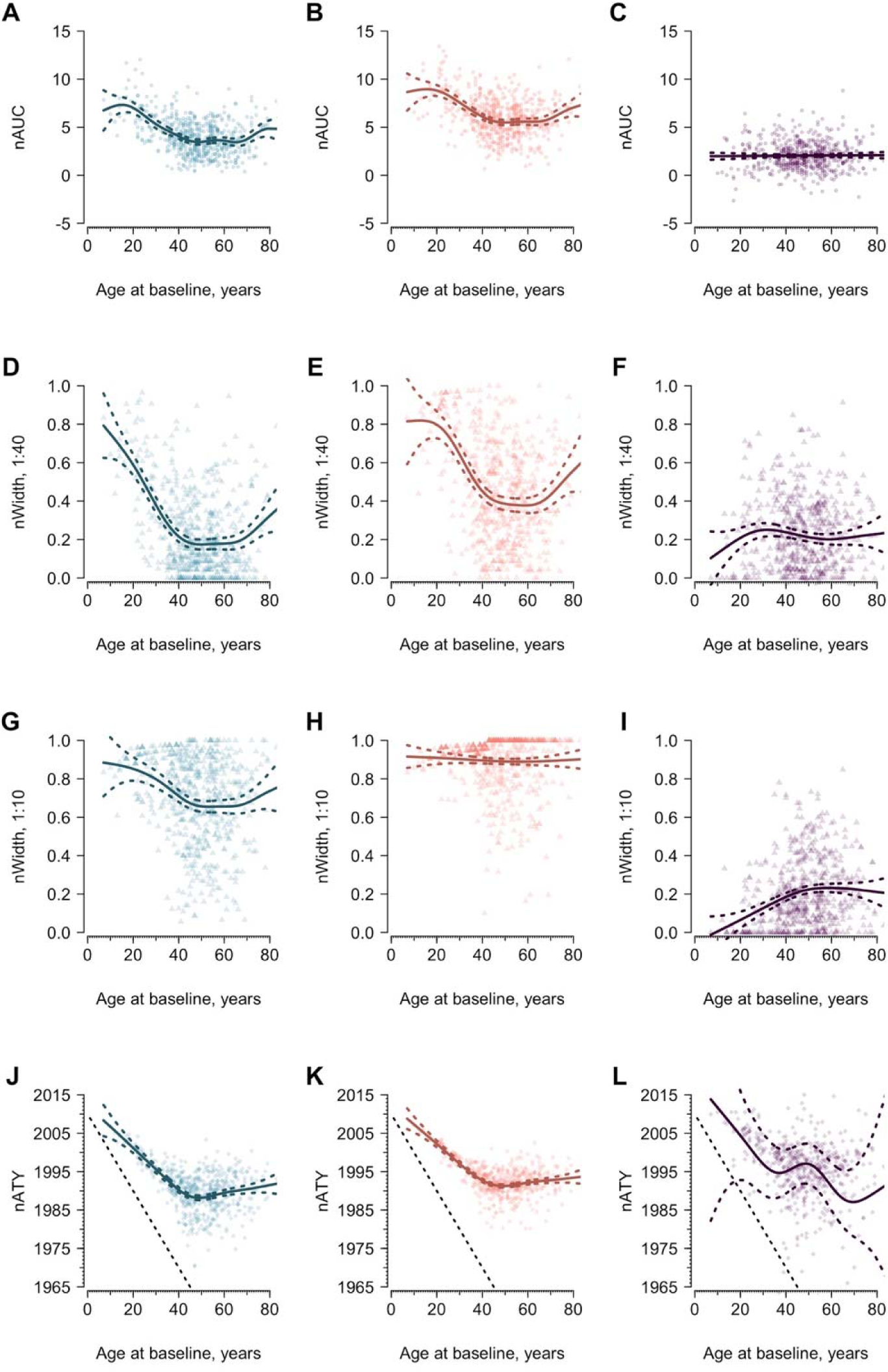
Area under the curves (AUC), average titer years (ATY) and width varying with age, excluding participants who self-reported had been vaccinated against influenza. Blue and red represent the AUC for the baseline and follow-up visit, respectively. Purple indicates the differences of indicators between the two visits. Solid lines are predictions from gam and the colored dashed lines represent the corresponding 95% confidence intervals. The sloping black dotted lines in panel J to L indicate the year of birth of participants. The dashed lines in panel J to L indicate the unweighted average isolation year of post-birth strains.

**Fig. S11.**
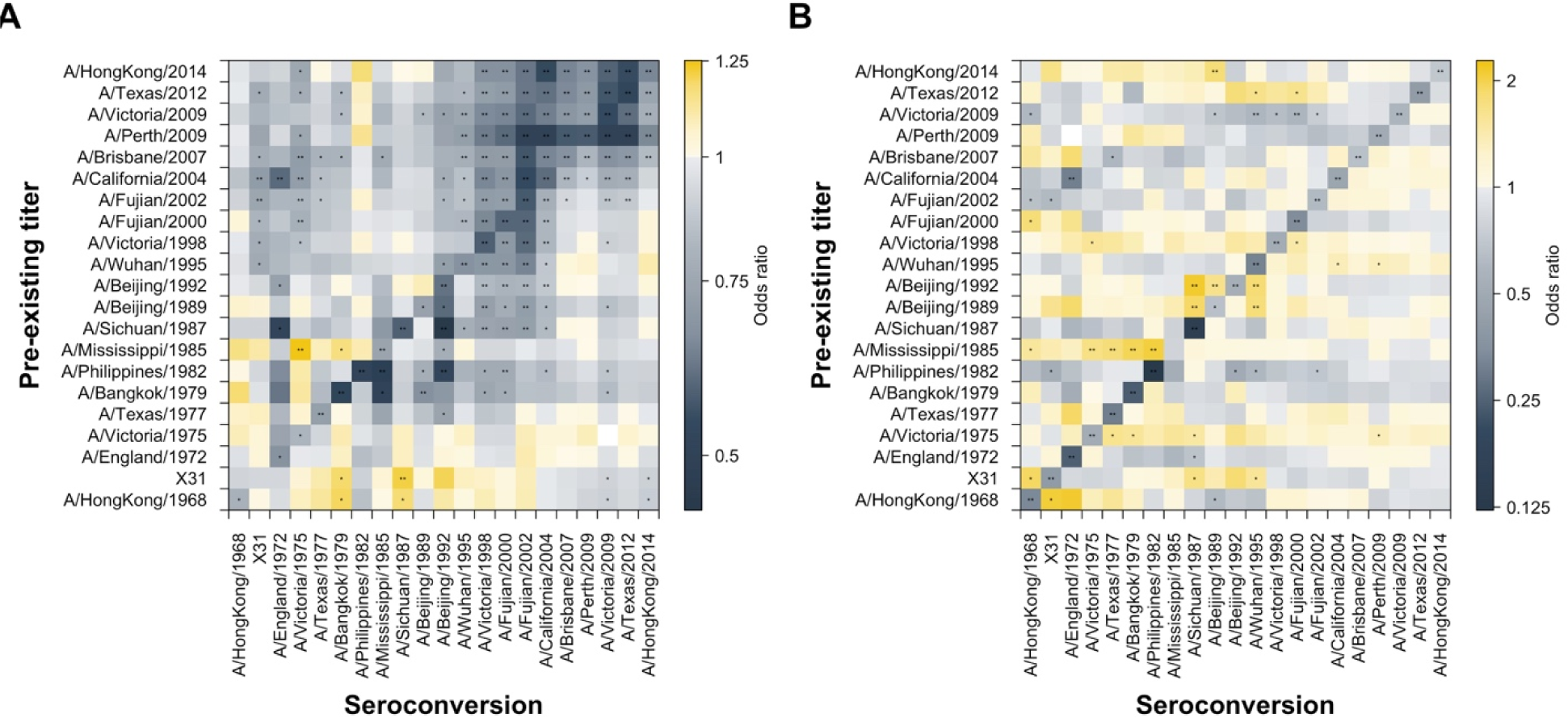
Association between pre-existing titer and seroconversion. (**A**) Univariable analysis of pre-existing titer on seroconversion. Coefficient was derived from univariable logistic regression of seroconversion to strain in x-axis on pre-existing titer to a strain listed in y-axis. Each cell represents an individual model. (**B**) Multivariable analysis of pre-existing titers on seroconversion. Coefficients were derived from multivariable logistic regression of seroconversion to a strain in x-axis on age at sampling and pre-existing titers to all strains listed in y-axis. Each column represents an individual model. Asterisks indicate p ≤ 0.01.

**Fig. S12.**
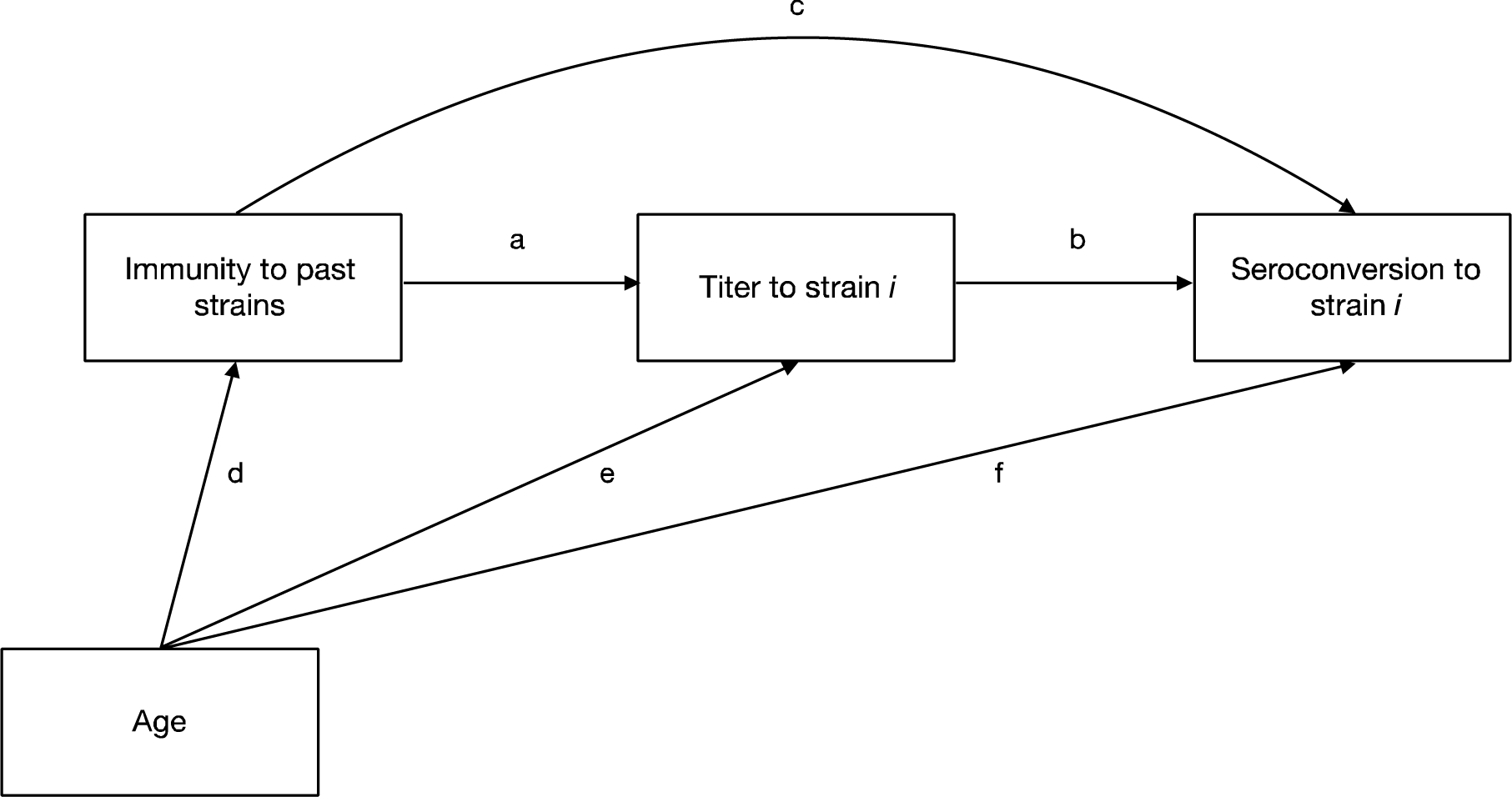
Directed acyclic graphs of hypothesized relations between immune responses to past strains, titer to recent strain and seroconversion to recent strain. Indirect effect (*path a path b*): immune responses to previous strains have positive association between titer to strain *i* due to cross-reactions (*path a*), which has a negative association with seroconversion to strain *i* (*path b*). Direct effect (*path c*): effect of immune responses on seroconversion to strain *i* that was not mediated by titer to strain *i*. Total effect (*path a path b* + *path c*): combination of indirect effect and direct effect. Confounding effect (*path d, e* and *f*).

**Fig. S13.**
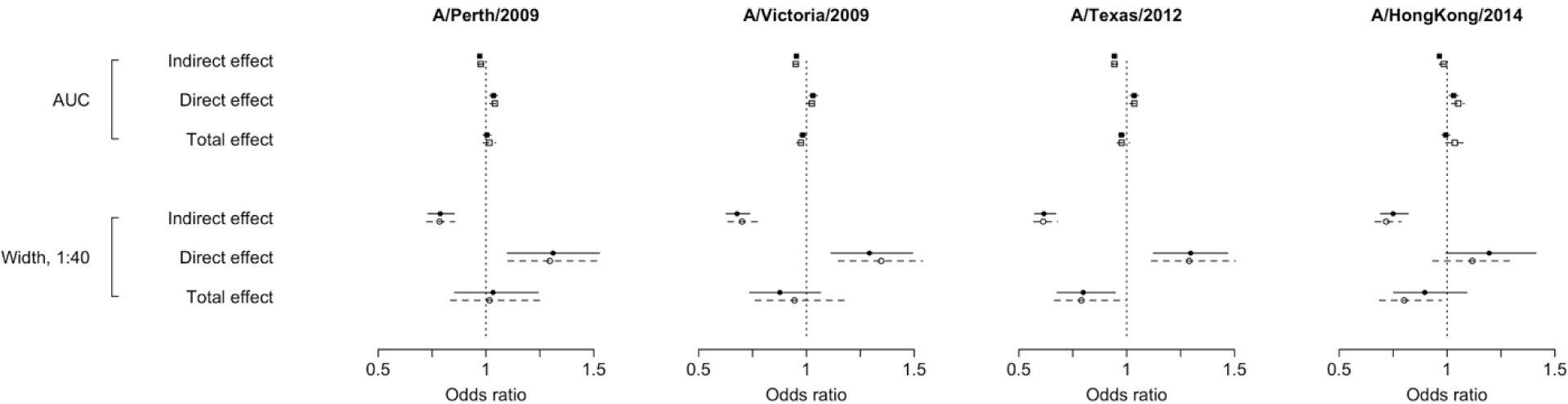
Mediation analysis of the effects of immune responses to previous strain on seroconversion to a recent strain. Solid lines and filled squares represent the estimates from mediation analysis that did not consider interactions. Dashed lines and open circles represent the estimates from mediation analysis that considered interactions.

**Fig. S14.**
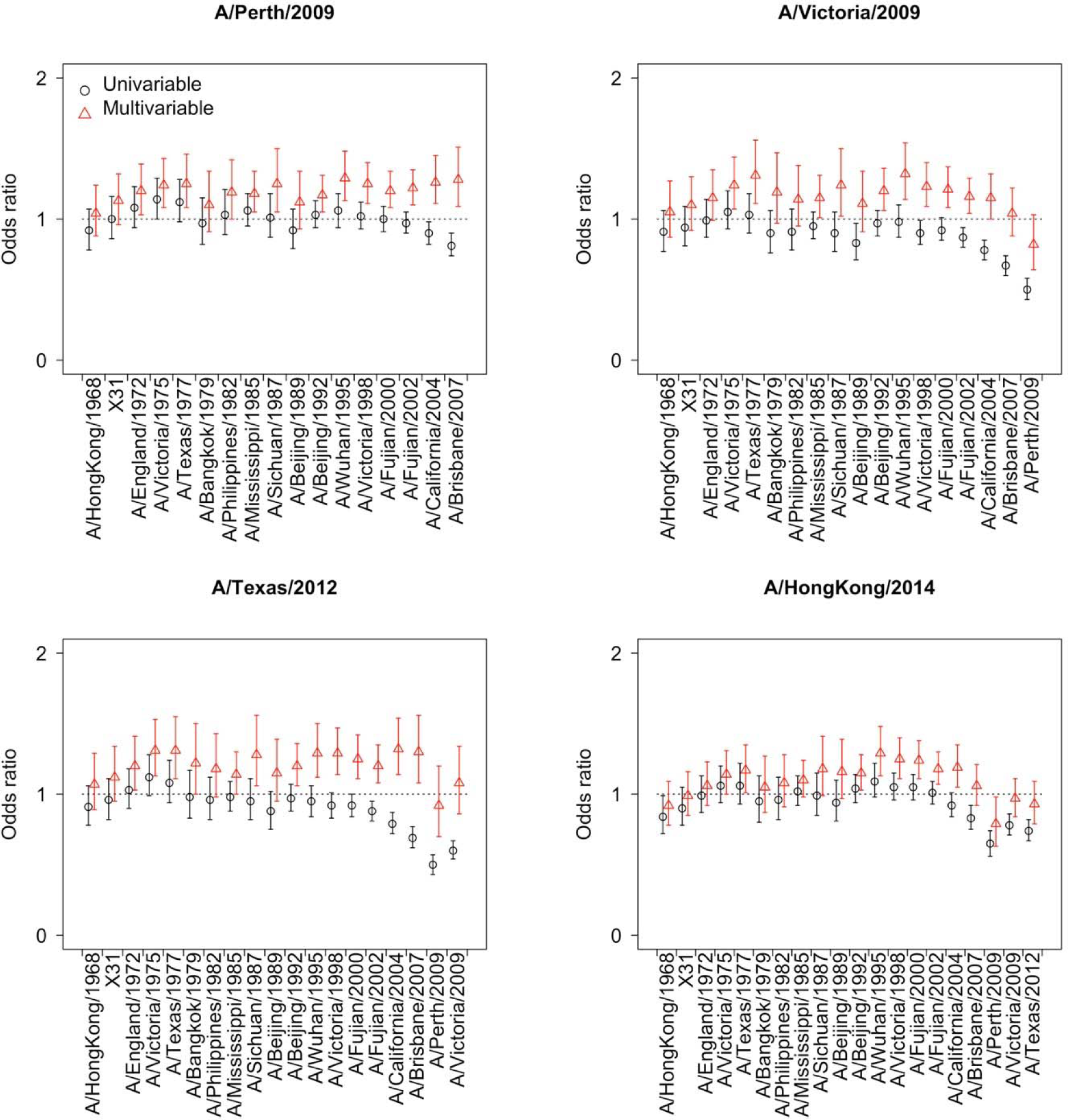
Association between pre-existing titer to individual strain and seroconversion to recent four strains. Univariable coefficient (black) is estimated from univariable logistic regression of seroconversion to strain i on pre-existing titer to the strain listed in x-axis. Multivariable coefficient (red) is estimated from multivariable logistic regression of seroconversion to a strain *i* on pre-existing titer to the strain listed in x-axis, adjusting for age at sampling and titer to strain *i* and *i-1*.

**Fig. S15.**
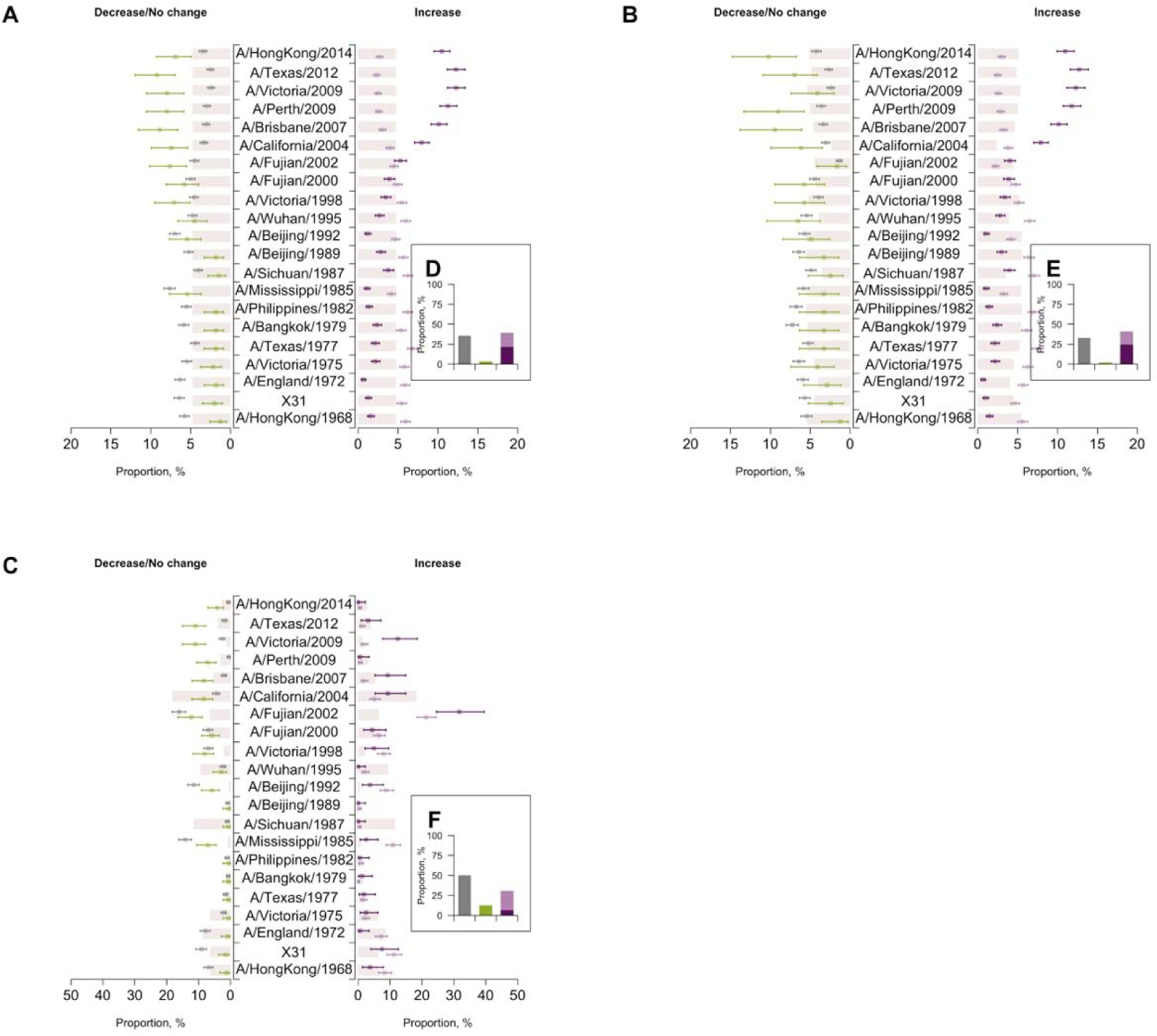
Distribution of H3N2 strains by changes in titers between two visits. We divided the examined data (i.e. all data, or pre-existing titer is greater or less than 1:80) on titer changes into four subgroups, i.e. decreased (green), unchanged (grey), any fold increase (light purple, including four-fold or more increase) and four-fold or more increase (dark purple). Colored points and lines represent the distribution of H3N2 strains within each subgroup. Colored bars represent the distribution of H3N2 strains regardless of titer changes for the examined data. (**A**) all data; (**B**) a subset contains pre-existing titers ≤1:40; (**C**) a subset contains pre-existing titers > 1:40. Insets **D** to **F** illustrate the distribution of changes in titers between two visits.

**Fig. S16.**
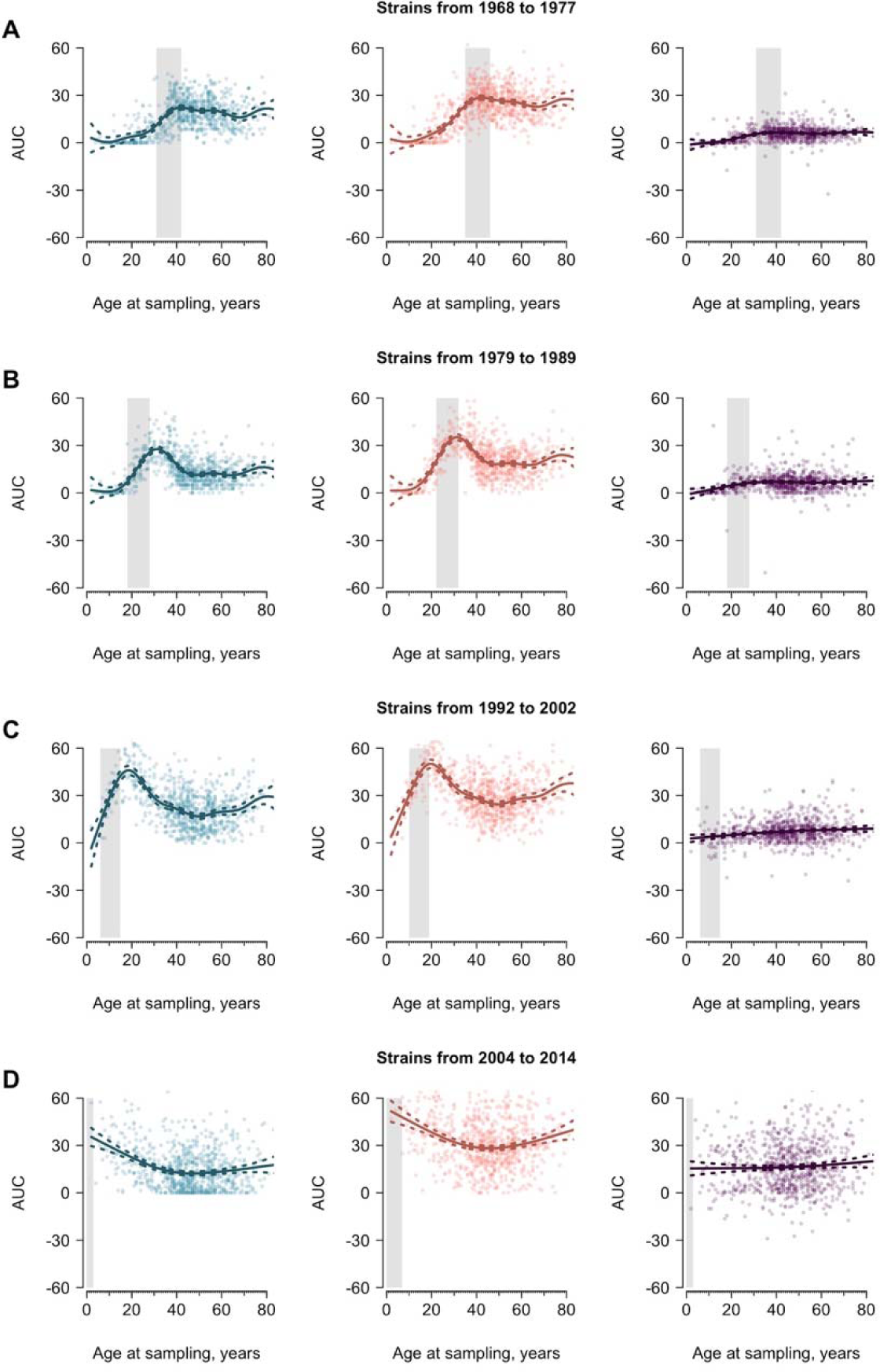
Average areas under curve (AUC) and differences in AUC between two visits varying with age, by strain groups. Blue and red represent the AUC for the baseline and follow-up visit, respectively. Purple indicates the differences of indicators AUC the two visits. Solid lines are predictions from gam and the colored dashed lines represent the corresponding 95% confidence intervals. Grey bars represent the age cohort when the participants were between 0 to 10 years when the examined strains were isolated. The results were calculated after stratifying by four strain groups, where we chronologically grouped the strains according to the year of isolation. (**A**) A/HongKong/1968, X31, A/England/1972, A/Victoria/1975 and A/Texas 1977. (**B**) A/Bangkok/1979, A/Philippines/1982, A/Mississippi/1985, A/Sichuan/1987, and A/Beijing/1989. (**C**) A/Beijing/1992, A/Wuhan/1995, A/Victoria/1998, A/Fujian/2000 and A/Fujian/2002. (**D**) A/California/2004, A/Brisbane/2007, A/Perth/2009, A/Victoria/2009, A/Texas/2012 and A/HongKong/2014.

**Fig. S17.**
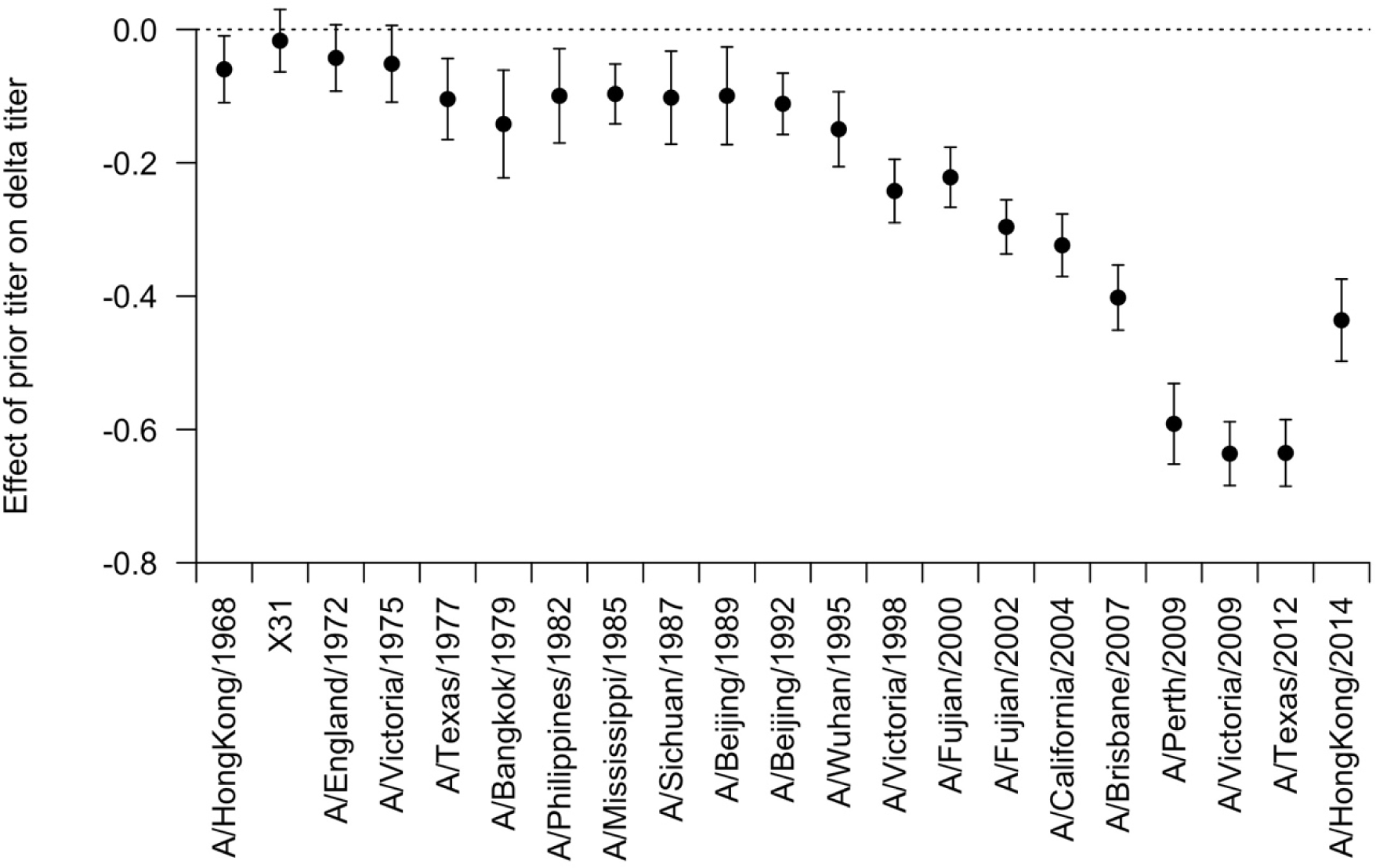
Effect of pre-existing HAI titer on differences in HAI titer between two visits varying from H3N2 strains. Linear regression models were fitted using prior titer to predict differences in HAI titer between two visits, with interaction term between prior titer and H3N2 strains and adjustment for strain and age at sampling.

**Fig. S18.**
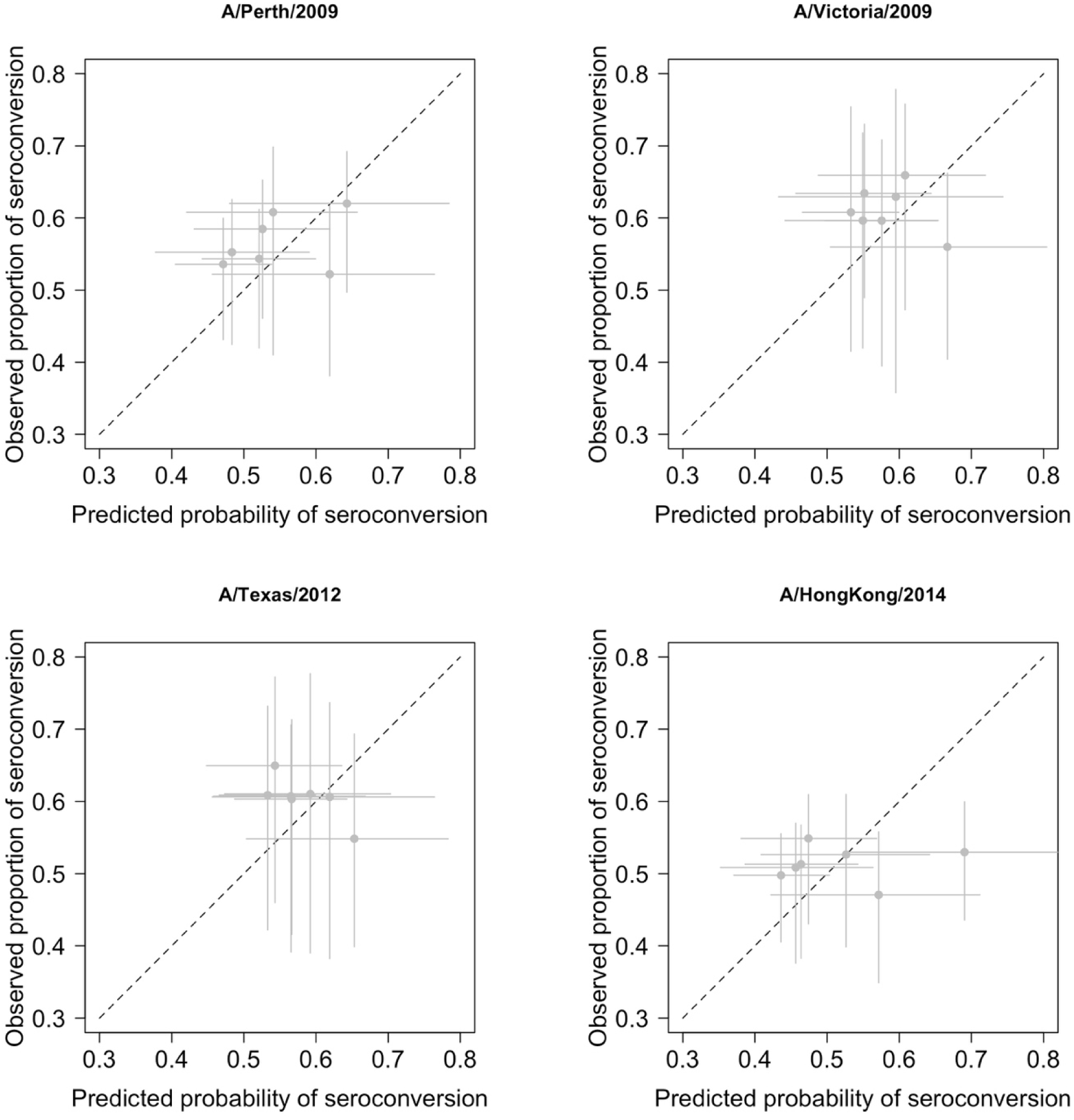
Predicted probability of seroconversion and observed proportion of seroconversion by age group. Models are fitted with a linear term on age, i.e. models used in Table 1. Age group was binned by 10 years. Horizontal lines represent the interquartile of predicted probability of seroconversion for the age group. Vertical lines represent 95% CI of the observed proportion of seroconversion derived from binomial distribution.

**Fig. S19.**
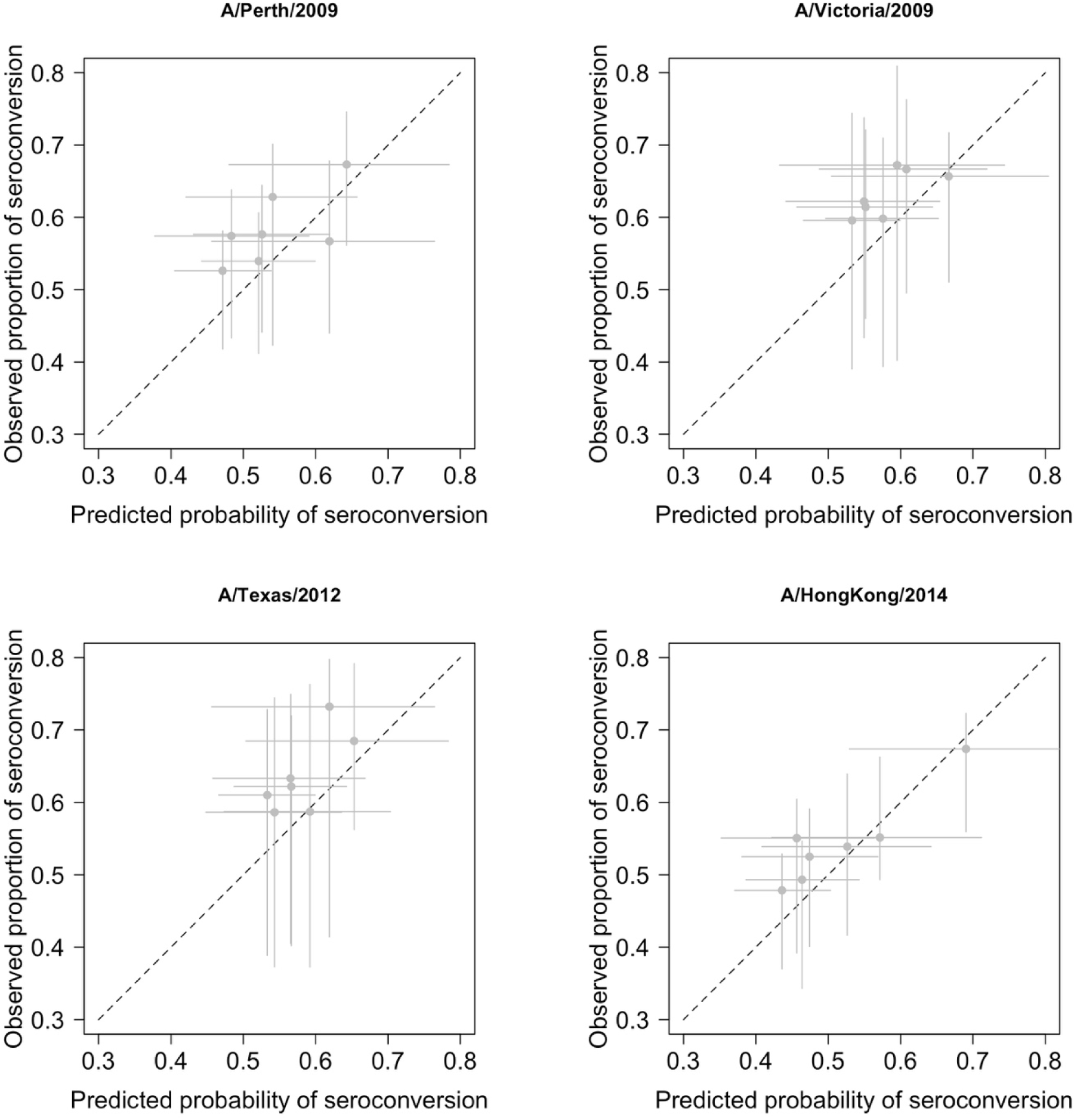
Predicted probability of seroconversion and observed proportion of seroconversion by age group, accounting for non-linear effect of age. Models are fitted with a spline term on age, i.e. models used in table S9. Age group was binned by 10 years. Horizontal lines represent the interquartile of predicted probability of seroconversion for the age group. Vertical lines represent 95% CI of the observed proportion of seroconversion derived from binomial distribution.

## Supplementary Tables

**Table S1.**
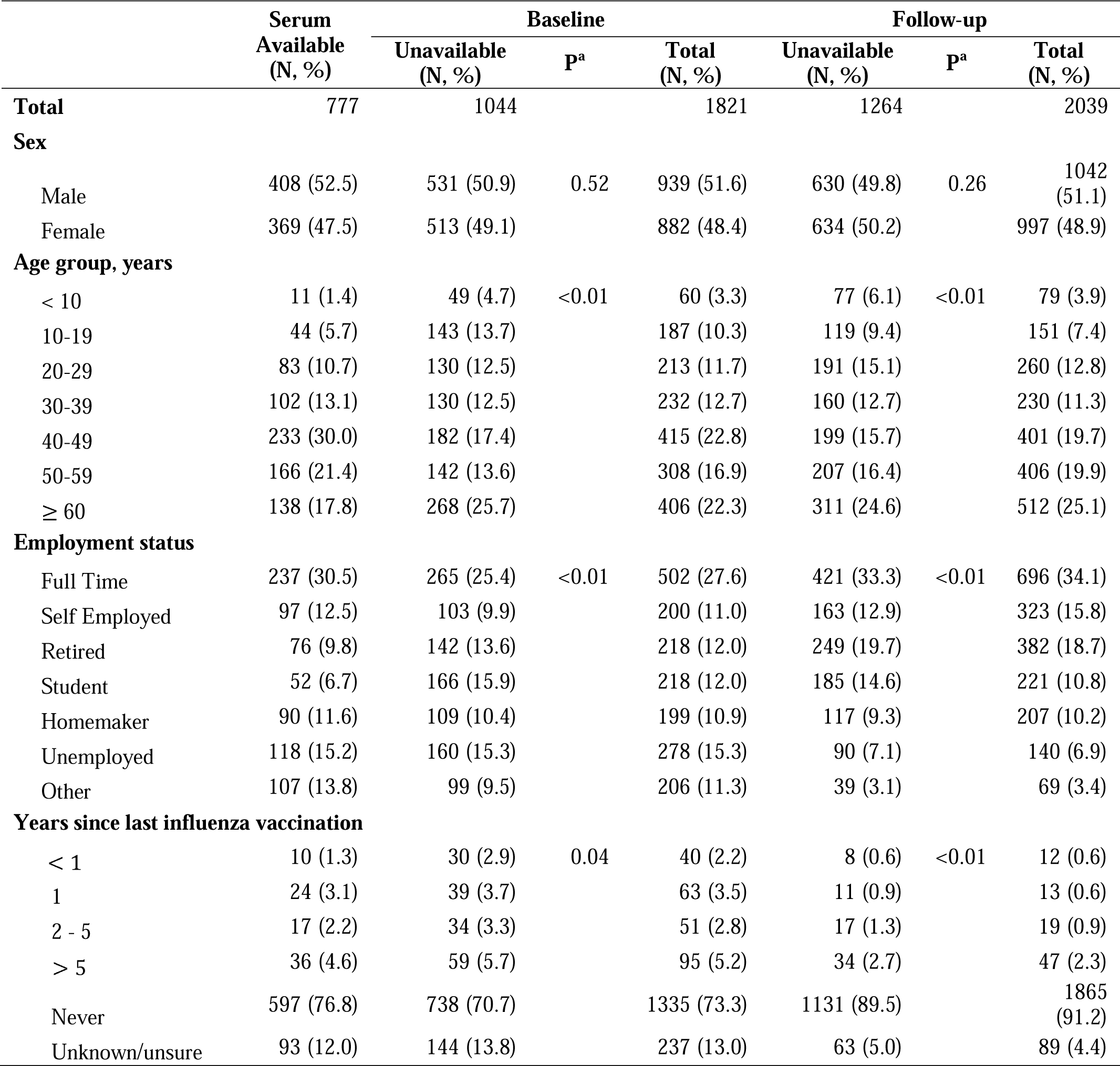
Comparison of demographic characteristics of participants.

**Table S2.** Geometric mean titer of tested H3N2 strains.

| Strain | All tested strains |  | Post-birth strains |  |
| --- | --- | --- | --- | --- |
|  | Baseline | Follow-up | Baseline | Follow-up |
| A/HongKong/1968 | 23.7 (22.0, 25.4) | 36.4 (33.7, 39.2) | 36.8 (34.2, 39.5) | 59.1 (55.2, 63.3) |
| X31 | 27.2 (25.2, 29.4) | 39.9 (36.7, 43.4) | 44.1 (41.2, 47.3) | 68.0 (63.3, 73.0) |
| A/England/1972 | 22.3 (20.7, 23.9) | 32.1 (29.8, 34.7) | 30.0 (27.9, 32.2) | 45.3 (42.2, 48.5) |
| A/Victoria/1975 | 12.1 (11.4, 12.9) | 19.4 (18.0, 20.7) | 14.3 (13.3, 15.4) | 24.1 (22.4, 26.0) |
| A/Texas/1977 | 13.1 (12.3, 13.9) | 22.0 (20.6, 23.5) | 14.1 (13.2, 15.0) | 24.9 (23.3, 26.6) |
| A/Bangkok/1979 | 7.7 (7.4, 8.1) | 12.3 (11.6, 12.9) | 8.0 (7.6, 8.4) | 13.3 (12.6, 14.1) |
| A/Philippines/1982 | 11.4 (10.8, 12.0) | 17.7 (16.8, 18.8) | 12.2 (11.5, 12.8) | 19.6 (18.5, 20.8) |
| A/Mississippi/1985 | 38.5 (35.5, 41.6) | 51.2 (47.1, 55.6) | 46.3 (42.9, 49.9) | 62.2 (57.6, 67.2) |
| A/Sichuan/1987 | 9.2 (8.7, 9.6) | 16.7 (15.7, 17.7) | 9.0 (8.5, 9.4) | 16.8 (15.8, 17.8) |
| A/Beijing/1989 | 8.5 (8.1, 8.9) | 14.0 (13.2, 14.8) | 8.6 (8.2, 9.0) | 14.5 (13.7, 15.4) |
| A/Beijing/1992 | 32.9 (30.5, 35.6) | 45.5 (42.2, 49.2) | 34.8 (32.2, 37.6) | 48.8 (45.3, 52.6) |
| A/Wuhan/1995 | 13.4 (12.5, 14.2) | 21.9 (20.5, 23.4) | 13.4 (12.5, 14.2) | 22.3 (20.9, 23.8) |
| A/Victoria/1998 | 27.7 (25.7, 29.9) | 46.8 (43.6, 50.2) | 27.6 (25.6, 29.7) | 46.7 (43.5, 50.1) |
| A/Fujian/2000 | 25.9 (23.9, 28.0) | 43.9 (40.7, 47.3) | 26.1 (24.1, 28.2) | 44.0 (40.8, 47.4) |
| A/Fujian/2002 | 71.8<br>(65.7, 78.3) | 133.1<br>(123.0, 144.1) | 72.1<br>(66.0, 78.7) | 133.7<br>(123.5, 144.7) |
| A/California/2004 | 20.6 (19.0, 22.2) | 47.1 (43.6, 50.9) | 20.6 (19.0, 22.2) | 47.1 (43.6, 50.8) |
| A/Brisbane/2007 | 14.3 (13.3, 15.4) | 40.0 (36.8, 43.4) | 14.3 (13.3, 15.4) | 39.9 (36.7, 43.3) |
| A/Perth/2009 | 9.1 (8.6, 9.7) | 30.9 (28.4, 33.5) | 9.1 (8.6, 9.7) | 30.9 (28.4, 33.5) |
| A/Victoria/2009 | 17.5 (16.3, 18.9) | 67.6 (62.3, 73.3) | 17.5 (16.3, 18.9) | 67.6 (62.3, 73.3) |
| A/Texas/2012 | 13.3 (12.4, 14.3) | 50.6 (46.5, 55.0) | 13.3 (12.4, 14.3) | 50.6 (46.5, 55.0) |
| A/HongKong/2014 | 9.6 (9.0, 10.2) | 28.1 (25.9, 30.4) | 9.6 (9.0, 10.2) | 28.1 (25.9, 30.4) |
| Overall | 17.1 (16.8, 17.4) | 33.0 (32.4, 33.6) | 18.3 (18.0, 18.7) | 36.9 (36.2, 37.5) |

**Table S3.** Associations between pre-existing immunity and seroconversion to four recent strains, using titers to all tested strains.

|  | A/Perth/2009 | A/Victoria/2009 | A/Texas/2012 | A/HongKong/2014 |
| --- | --- | --- | --- | --- |
| **Model 1** |  |  |  |  |
| Age at sampling | 1.00 (0.99, 1.01) | 0.99 (0.98, 1.00) | 0.99 (0.98, 1.00)* | 1.00 (0.99, 1.01) |
| Titer to strain *i*<sup>a</sup> | 0.43 (0.34, 0.54)* | 0.51 (0.42, 0.60)* | 0.49 (0.38, 0.61)* | 0.70 (0.56, 0.85)* |
| Titer to strain *i-l*<sup>a</sup> | 1.30 (1.10, 1.55)* | 1.03 (0.87, 1.22) | 1.06 (0.85, 1.32) | 0.93 (0.79, 1.09) |
| Deviance explained | 7.5% | 12.9% | 12.6% | 4.0% |
| **Model 2** |  |  |  |  |
| Age at sampling | 1.00 (0.99, 1.01) | 0.99 (0.98, 1.00) | 0.99 (0.98, 1.00) | 1.00 (0.99, 1.01) |
| Titer to strain *i* | 0.43 (0.34, 0.54)* | 0.49 (0.41, 0.58)* | 0.47 (0.37, 0.60)* | 0.68 (0.55, 0.84)* |
| Titer to strain *i-l* | 1.26 (1.06, 1.50)* | 1.02 (0.86, 1.21) | 1.00 (0.80, 1.26) | 0.89 (0.75, 1.05) |
| AUC<sup>b</sup> | 1.01 (1.00, 1.01) | 1.01 (1.00, 1.01) | 1.01 (1.00, 1.01)* | 1.01 (1.00, 1.01) |
| Deviance explained | 7.8% | 13.3% | 13.0% | 4.4% |
| **Model 3** |  |  |  |  |
| Age at sampling | 1.00 (0.99, 1.01) | 0.99 (0.98, 1.00) | 0.99 (0.98, 1.00) | 1.00 (0.99, 1.01) |
| Titer to strain *i* | 0.43 (0.34, 0.54)* | 0.49 (0.41, 0.58)* | 0.47 (0.37, 0.60)* | 0.68 (0.55, 0.84)* |
| Titer to strain *i-l* | 1.26 (1.06, 1.50)* | 1.01 (0.85, 1.19) | 1.01 (0.80, 1.26) | 0.90 (0.76, 1.07) |
| Width, cut off 1:40<sup>b</sup> | 2.86 (0.99, 8.40) | 3.85 (1.24, 12.31)* | 4.15 (1.40, 12.63)* | 2.15 (0.80, 5.80) |
| Deviance explained | 7.8% | 13.4% | 13.2% | 4.2% |

<sup>a</sup> Strain *i* refers to the strain that was examined for seroconversion, and strain *i-l* refers to the most recent strain isolated prior to strain *i*. E.g. when using seroconversion to A/Perth/2009 as outcome, strain *i* and *i-l* will be A/Perth/2009 and A/Brisbane/2007, respectively.
<sup>b</sup> Metrics were calculated using titers to strains isolated after 1968 and before the year that strain *i* was isolated, regardless of whether the strain circulated before or after the birth of the participant.

**Table S4.**
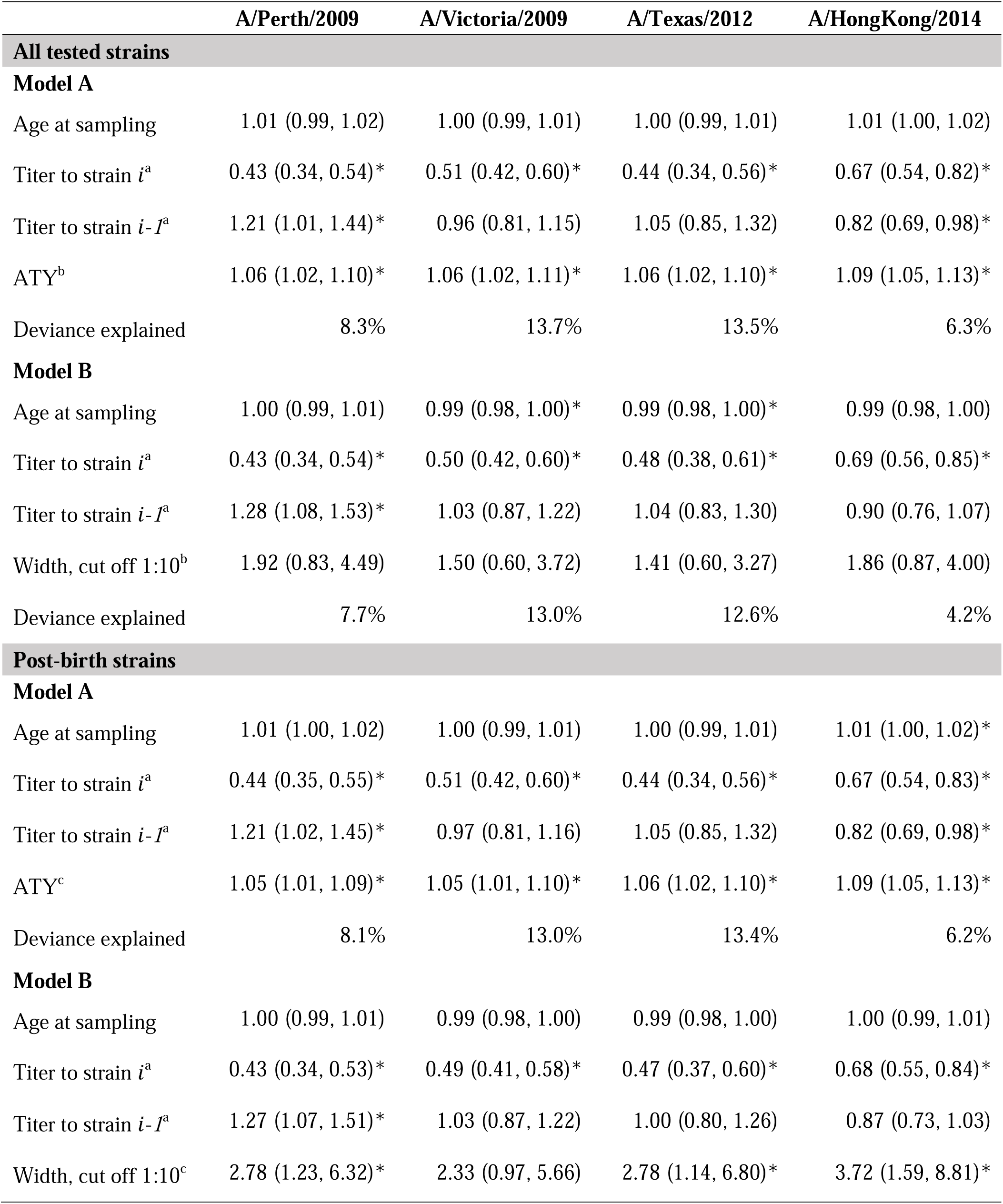
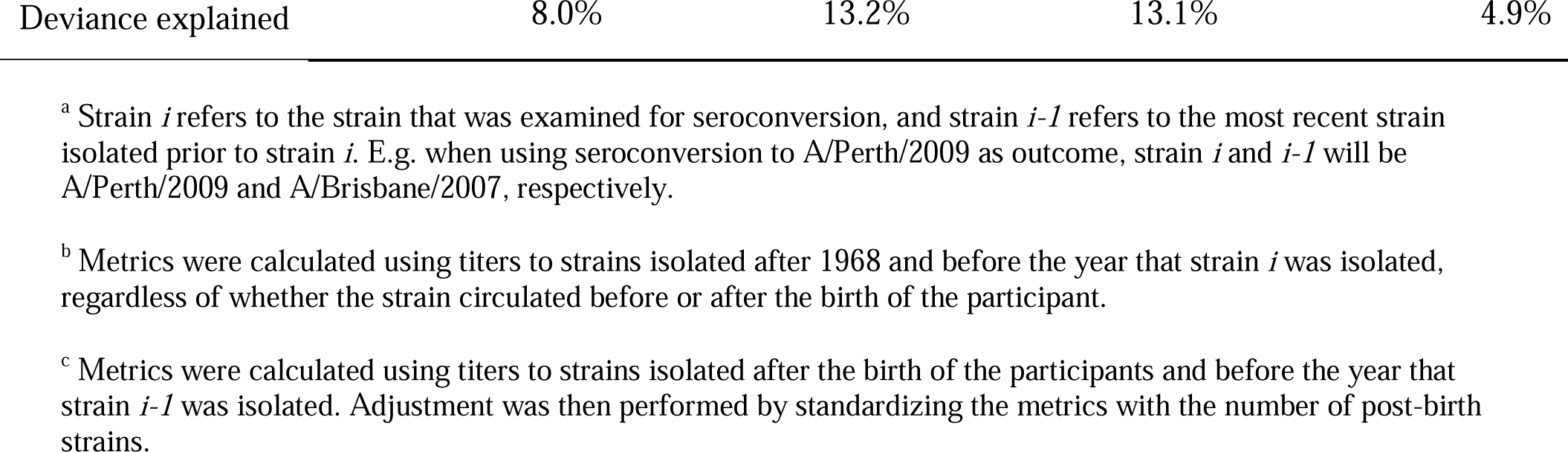
Associations between pre-existing average titer year or width above detectable threshold and seroconversion to four recent strains.

**Table S5.**
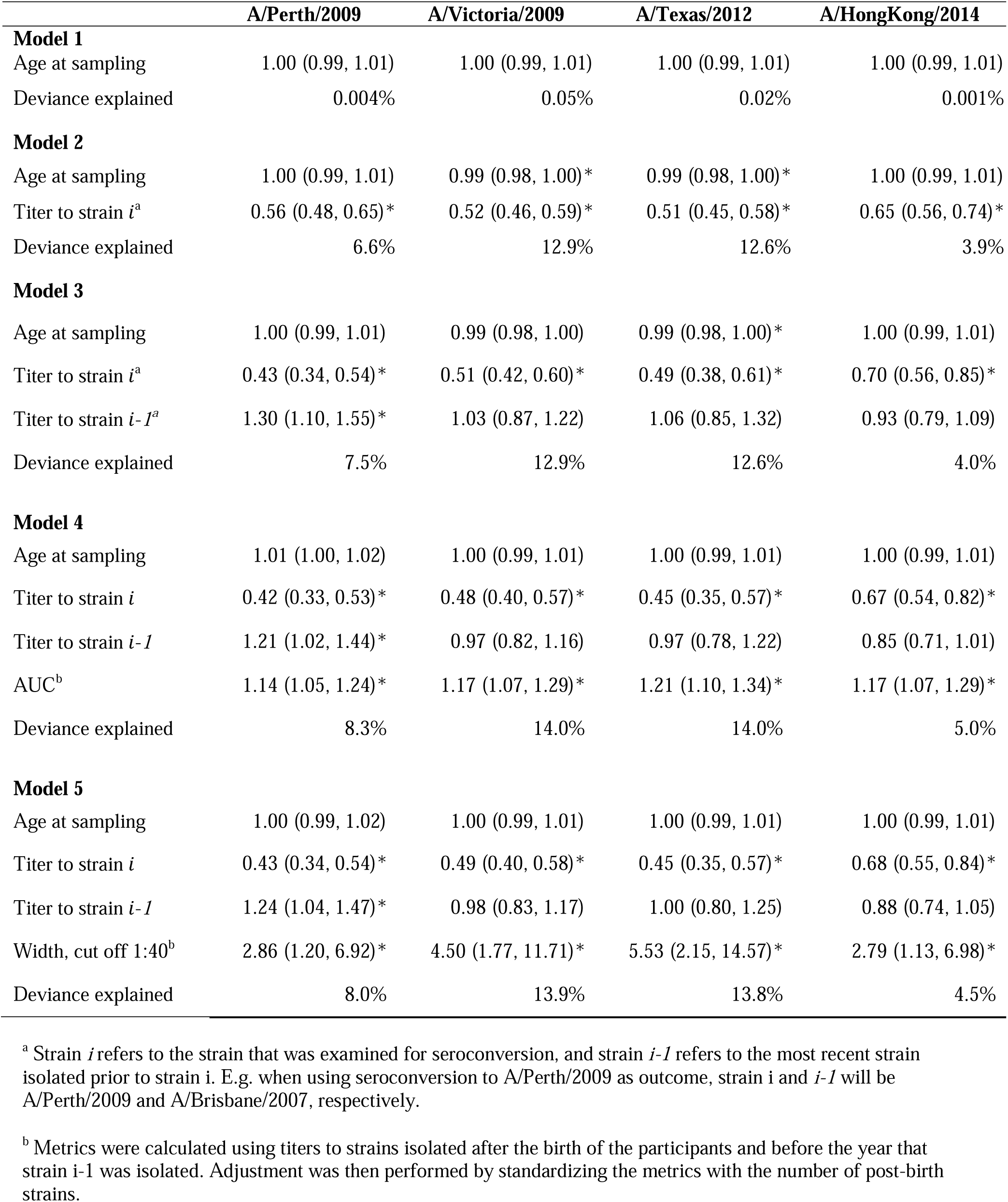
Associations between pre-existing immunity and seroconversion to four recent strains.

**Table S6.** Associations between pre-existing immunity and seroconversion to four recent strains after removing pre-existing titers to outcome strains.

|  | A/Perth/2009 | A/Victoria/2009 | A/Texas/2012 | A/HongKong/2014 |
| --- | --- | --- | --- | --- |
| **Areas under curves (AUC)** |  |  |  |  |
| **Model 1** |  |  |  |  |
| Age at sampling | 1.01 (1.00, 1.02) | 1.00 (0.99, 1.01) | 1.00 (0.99, 1.01) | 1.00 (0.99, 1.01) |
| Titer to strain $i^a$ | 0.42 (0.33, 0.53)* | 0.48 (0.40, 0.57)* | 0.45 (0.35, 0.57)* | 0.67 (0.54, 0.82)* |
| Titer to strain $i-I^a$ | 1.21 (1.02, 1.44)* | 0.97 (0.82, 1.16) | 0.97 (0.78, 1.22) | 0.85 (0.71, 1.01) |
| AUC<sup>b</sup> | 1.14 (1.05, 1.24)* | 1.17 (1.07, 1.29)* | 1.21 (1.10, 1.34)* | 1.17 (1.07, 1.29)* |
| Deviance explained | 8.3% | 14.0% | 14.0% | 5.0% |
| **Model 2** |  |  |  |  |
| Age at sampling | 1.00 (0.99, 1.01) | 0.99 (0.98, 1.00) | 1.00 (0.99, 1.01) | 1.00 (0.99, 1.01) |
| Titer to strain $i-I^a$ | 0.75 (0.67, 0.84)* | 0.62 (0.54, 0.70)* | 0.54 (0.47, 0.62)* | 0.68 (0.59, 0.77)* |
| AUC<sup>b</sup> | 1.11 (1.03, 1.21)* | 1.07 (0.98, 1.16) | 1.14 (1.04, 1.25)* | 1.15 (1.05, 1.27)* |
| Deviance explained | 2.3% | 6.5% | 9.7% | 3.6% |
| **Model 3** |  |  |  |  |
| Age at sampling | 1.00 (0.99, 1.01) | 0.99 (0.98, 1.00) | 0.99 (0.98, 1.00) | 1.00 (0.99, 1.01) |
| AUC<sup>b</sup> | 1.02 (0.95, 1.10) | 0.93 (0.86, 1.00)* | 0.90 (0.83, 0.96)* | 0.98 (0.91, 1.06) |
| Deviance explained | 0% | 0.5% | 0.9% | 0% |
| **Width, cut off 1:40** |  |  |  |  |
| **Model 1** |  |  |  |  |
| Age at sampling | 1.00 (0.99, 1.02) | 1.00 (0.99, 1.01) | 1.00 (0.99, 1.01) | 1.00 (0.99, 1.01) |
| Titer to strain $i^a$ | 0.43 (0.34, 0.54)* | 0.49 (0.40, 0.58)* | 0.45 (0.35, 0.57)* | 0.68 (0.55, 0.84)* |
| Titer to strain $i-I^a$ | 1.24 (1.04, 1.47)* | 0.98 (0.83, 1.17) | 1.00 (0.80, 1.25) | 0.88 (0.74, 1.05) |
| Width, cut off 1:40<sup>b</sup> | 2.86 (1.20, 6.92)* | 4.50 (1.77, 11.71)* | 5.53 (2.15, 14.57)* | 2.79 (1.13, 6.98)* |
| Deviance explained | 8.0% | 13.9% | 13.8% | 4.5% |
| **Model 2** |  |  |  |  |
| Age at sampling | 1.00 (0.99, 1.01) | 0.99 (0.98, 1.00) | 1.00 (0.99, 1.01) | 1.00 (0.99, 1.01) |
| Titer to strain $i-I^a$ | 0.77 (0.68, 0.86)* | 0.62 (0.54, 0.70)* | 0.55 (0.48, 0.63)* | 0.71 (0.62, 0.80)* |
| Width, cut off 1:40<sup>b</sup> | 2.48 (1.09, 5.72)* | 2.16 (0.92, 5.18) | 3.19 (1.31, 7.89)* | 2.35 (0.96, 5.79) |
| Deviance explained | 2.1% | 6.6% | 9.6% | 3.2% |
| **Model 3** |  |  |  |  |
| Age at sampling | 1.00 (0.99, 1.01) | 0.99 (0.98, 1.00) | 0.99 (0.98, 1.00) | 1.00 (0.99, 1.01) |
| Width, cut off 1:40<sup>b</sup> | 1.17 (0.55, 2.48) | 0.57 (0.27, 1.22) | 0.40 (0.19, 0.83)* | 0.68 (0.32, 1.46) |
| Deviance explained | 0% | 0.2% | 0.6% | 0.1% |

<sup>a</sup> Strain $i$ refers to the strain that was examined for seroconversion, and strain $i-1$ refers to the most recent strain isolated prior to strain $i$ . E.g. when using seroconversion to A/Perth/2009 as outcome, strain $i$ and $i-1$ will be A/Perth/2009 and A/Brisbane/2007, respectively.
<sup>b</sup> Metrics were calculated using titers to strains isolated after the birth of the participants and before the year that strain $i-1$ was isolated. Adjustment was then performed by standardizing the metrics with the number of post-birth strains.

**Table S7.** Associations between and pre-existing immunity and seroconversion to four recent strains, considering the non-linear impact of age.

|  | A/Perth/2009 | A/Victoria/2009 | A/Texas/2012 | A/HongKong/2014 |
| --- | --- | --- | --- | --- |
| **Model 1** |  |  |  |  |
| Titer to strain *i*<sup>a</sup> | 0.44 (0.35, 0.55)* | 0.52 (0.43, 0.62)* | 0.45 (0.35, 0.58)* | 0.69 (0.56, 0.85)* |
| Titer to strain *i-I*<sup>a</sup> | 1.25 (1.05, 1.48)* | 0.98 (0.83, 1.16) | 1.09 (0.87, 1.36) | 0.90 (0.76, 1.06) |
| Deviance explained | 8.2% | 13.7% | 14.3% | 5.8% |
| **Model 2** |  |  |  |  |
| Titer to strain *i* | 0.43 (0.34, 0.54)* | 0.49 (0.40, 0.58)* | 0.41 (0.32, 0.53)* | 0.67 (0.54, 0.83)* |
| Titer to strain *i-I* | 1.18 (0.99, 1.41) | 0.94 (0.79, 1.12) | 0.99 (0.79, 1.26) | 0.84 (0.70, 1.00) |
| AUC<sup>b</sup> | 1.12 (1.03, 1.23)* | 1.16 (1.05, 1.27)* | 1.21 (1.08, 1.36)* | 1.13 (1.02, 1.25)* |
| Deviance explained | 8.8% | 14.5% | 16.4% | 6.3% |
| **Model 3** |  |  |  |  |
| Titer to strain *i* | 0.43 (0.35, 0.55)* | 0.50 (0.41, 0.59)* | 0.43 (0.34, 0.55)* | 0.68 (0.55, 0.84)* |
| Titer to strain *i-I* | 1.21 (1.01, 1.44)* | 0.94 (0.79, 1.13) | 1.03 (0.82, 1.30) | 0.88 (0.74, 1.04) |
| Width, cut off 1:40<sup>b</sup> | 2.38 (0.96, 5.89) | 3.84 (1.44, 10.24)* | 3.72 (1.32, 10.51)* | 1.68 (0.64, 4.43) |
| Deviance explained | 8.5% | 14.4% | 15.1% | 5.9% |

<sup>a</sup> Strain *i* refers to the strain that was examined for seroconversion, and strain *i-I* refers to the most recent strain isolated prior to strain *i*. E.g. when using seroconversion to A/Perth/2009 as outcome, strain *i* and *i-I* will be A/Perth/2009 and A/Brisbane/2007, respectively.
<sup>b</sup> Metrics were calculated using titers to strains isolated after the birth of the participants and before the year that strain *i-I* was isolated. Adjustment was then performed by standardizing the metrics with the number of post-birth strains.

**Table S8.** Comparison of demographic characteristics of participants who self-reported to have not been vaccinated against influenza.

|  | All (N, %) | Never vaccinated<br>(N, %) | P<sup>a</sup> |
| --- | --- | --- | --- |
| **Total** | 777 | 573 |  |
| **Sex** |  |  |  |
| Male | 408 (52.5) | 283 (49.4) | 0.28 |
| Female | 369 (47.5) | 290 (50.6) |  |
| **Age group, years** |  |  |  |
| < 10 | 11 (1.4) | 1 (0.2) | <0.01 |
| 10-19 | 44 (5.7) | 13 (2.3) |  |
| 20-29 | 83 (10.7) | 52 (9.1) |  |
| 30-39 | 102 (13.1) | 71 (12.4) |  |
| 40-49 | 233 (30.0) | 178 (31.1) |  |
| 50-59 | 166 (21.4) | 145 (25.3) |  |
| ≥ 60 | 138 (17.8) | 113 (19.7) |  |
| **Employment status** |  |  |  |
| Full Time | 237 (30.5) | 174 (30.4) | 0.05 |
| Self Employed | 97 (12.5) | 80 (14.0) |  |
| Retired | 76 (9.8) | 60 (10.5) |  |
| Student | 52 (6.7) | 15 (2.6) |  |
| Homemaker | 90 (11.6) | 74 (12.9) |  |
| Unemployed | 118 (15.2) | 85 (14.8) |  |
| Other | 107 (13.8) | 85 (14.8) |  |

<sup>a</sup> P-value was derived from Chi-square test or Fisher exact test. We compared the distributions between individuals provided or not provided blood samples for each visit separately.

**Table S9.**
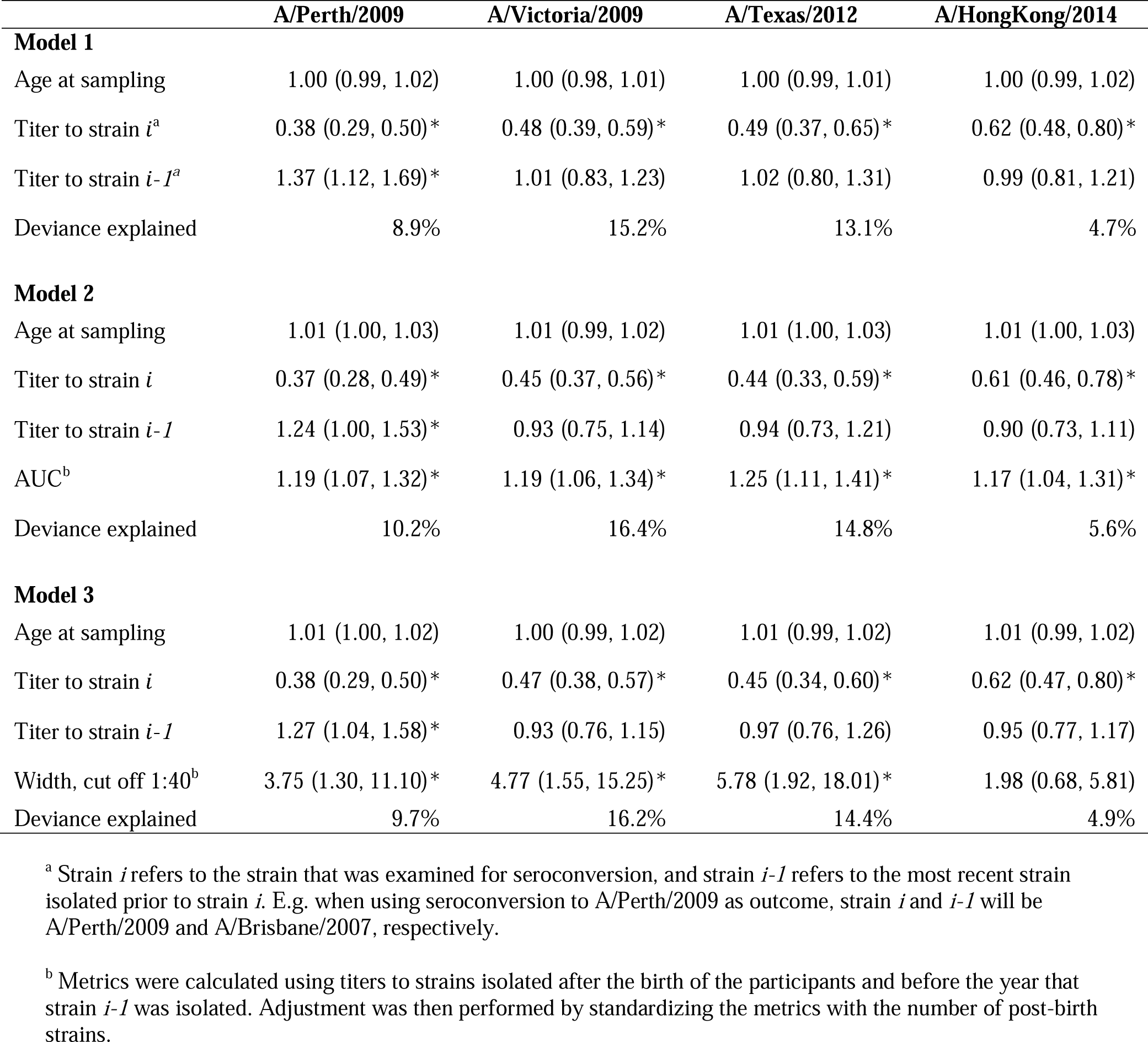
Associations between and pre-existing immunity and seroconversion to four recent strains, participants who reported never had been vaccinated against influenza.

**Table S10.**
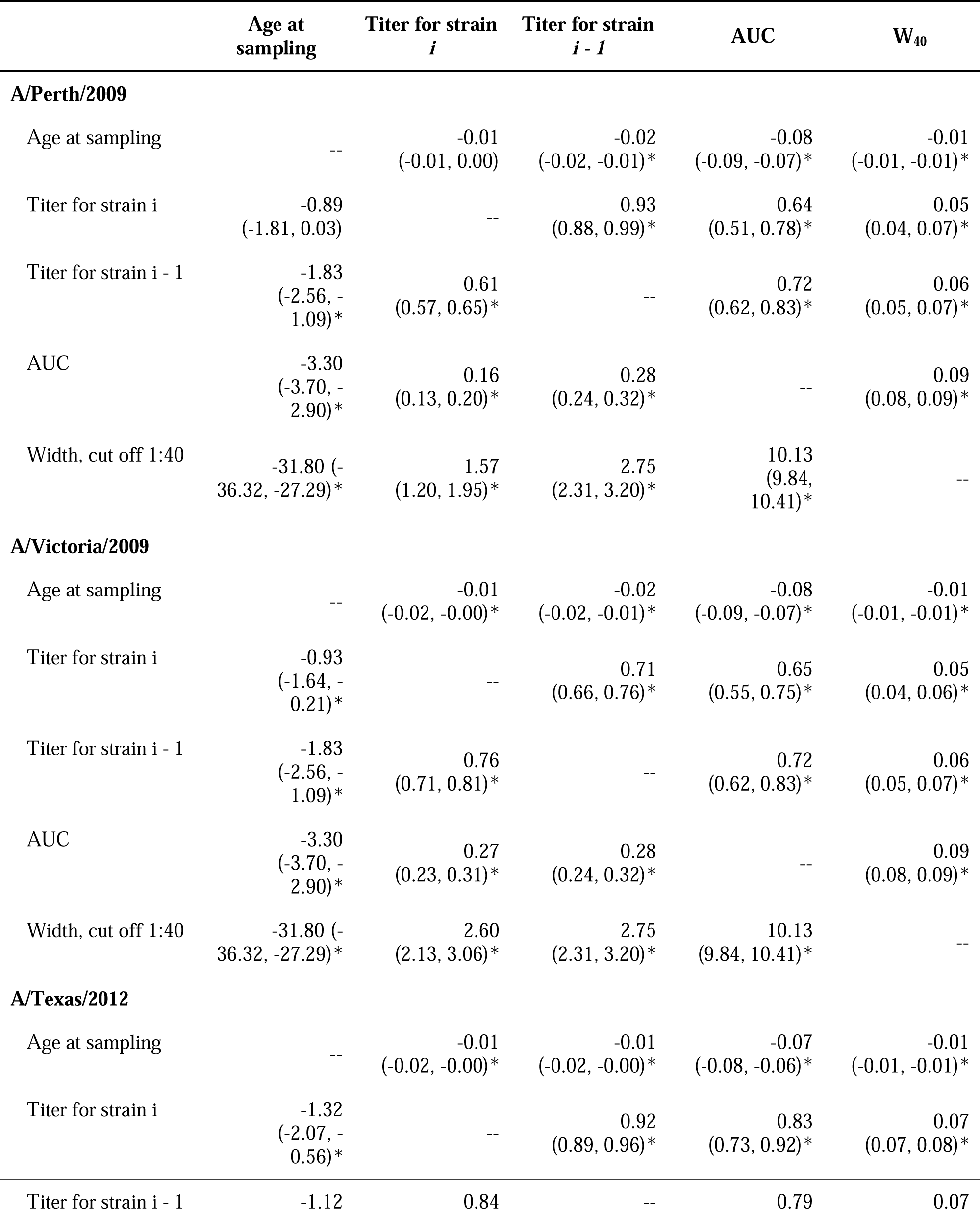
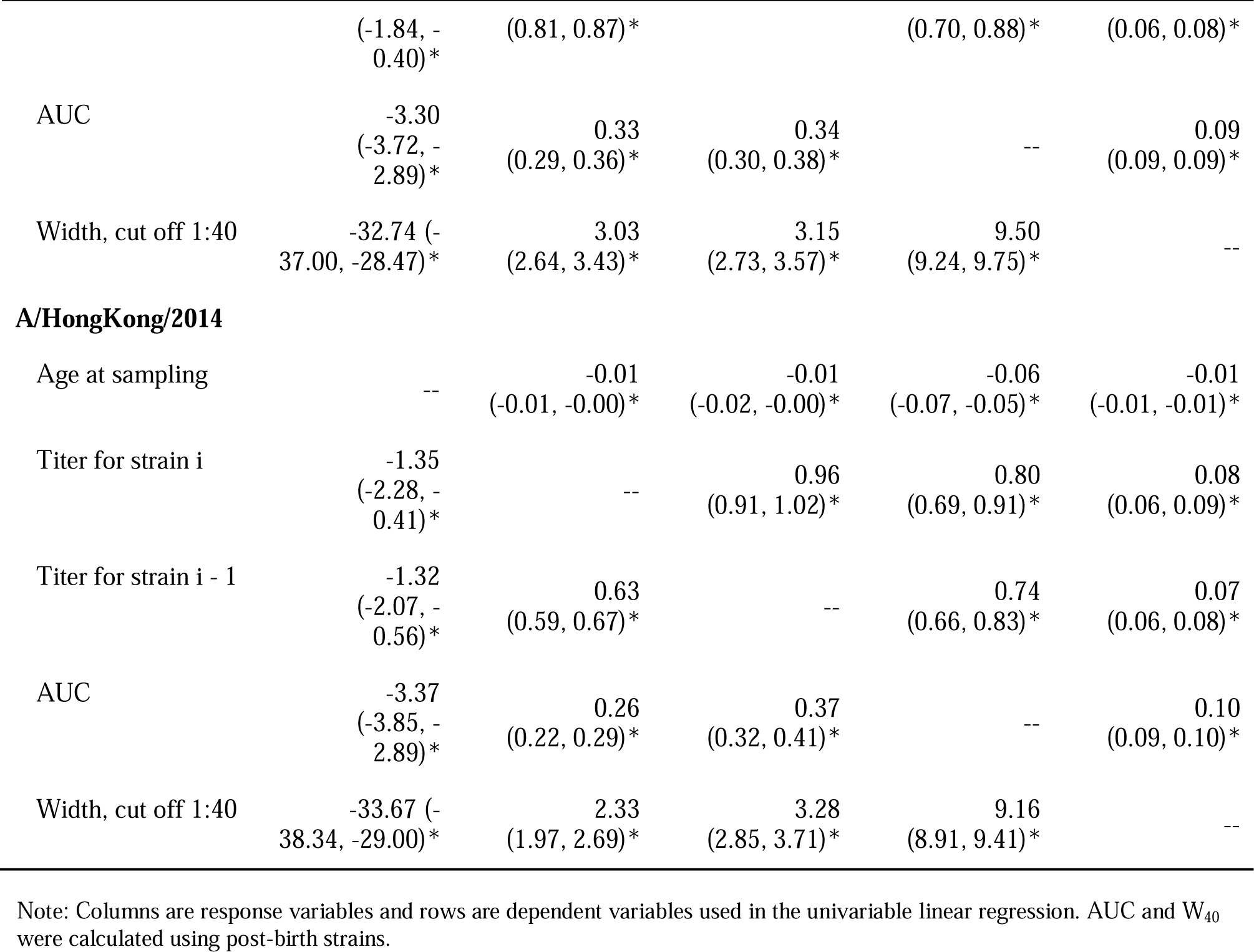
Univariable analysis of predictors used to assess the association between pre-existing immunity and seroconversion to four recent strains.

**Table S11.** Univariable logistic regressions of seroconversion to four recent strains on age and pre-existing immunity.

|  | **A/Perth/2009** | **A/Victoria/2009** | **A/Texas/2012** | **A/HongKong/2014** |
| --- | --- | --- | --- | --- |
| Age at sampling | 1.00 (0.99, 1.01) | 1.00 (0.99, 1.01) | 1.00 (0.99, 1.01) | 1.00 (0.99, 1.01) |
| Titer for strain $i$ | 0.56 (0.48, 0.65)* | 0.53 (0.47, 0.60)* | 0.52 (0.46, 0.59)* | 0.65 (0.57, 0.75)* |
| Titer for strain $i - I$ | 0.81 (0.73, 0.90)* | 0.66 (0.59, 0.74)* | 0.60 (0.54, 0.67)* | 0.75 (0.67, 0.83)* |
| AUC, post-birth strains | 1.02 (0.96, 1.08) | 0.96 (0.90, 1.02) | 0.93 (0.87, 0.99)* | 0.99 (0.92, 1.06) |
| ATY, post-birth strains | 1.00 (0.95, 1.04) | 0.94 (0.90, 0.99)* | 0.91 (0.88, 0.95)* | 1.00 (0.97, 1.04) |
| $W_{10}$, post-birth strains | 1.45 (0.68, 3.08) | 0.47 (0.22, 1.01) | 0.36 (0.17, 0.75)* | 1.13 (0.54, 2.38) |
| $W_{40}$, post-birth strains | 1.17 (0.60, 2.29) | 0.72 (0.37, 1.41) | 0.53 (0.28, 1.00)* | 0.75 (0.38, 1.48) |
| AUC, all strains | 1.00 (0.99, 1.01) | 0.99 (0.99, 1.00)* | 0.99 (0.99, 1.00)* | 1.00 (0.99, 1.00) |
| ATY, all strains | 1.02 (0.99, 1.05) | 1.00 (0.97, 1.03) | 0.97 (0.95, 1.00)* | 1.02 (0.99, 1.05) |
| $W_{10}$, all strains | 1.11 (0.51, 2.44) | 0.35 (0.15, 0.77)* | 0.32 (0.15, 0.64)* | 0.83 (0.41, 1.65) |
| $W_{40}$, all strains | 1.07 (0.43, 2.66) | 0.42 (0.17, 1.05) | 0.33 (0.14, 0.75)* | 0.57 (0.25, 1.30) |

**Table S12.**
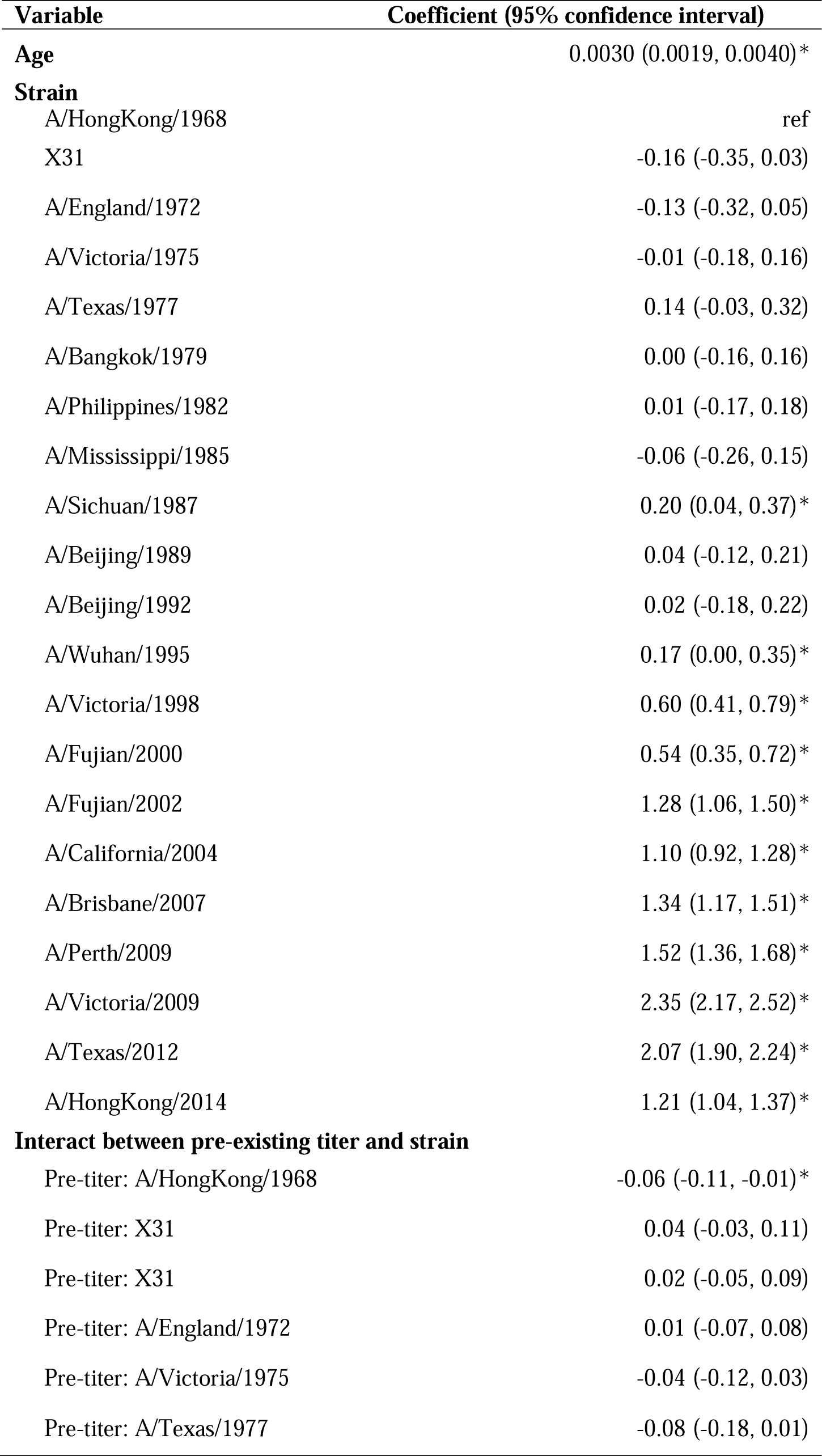
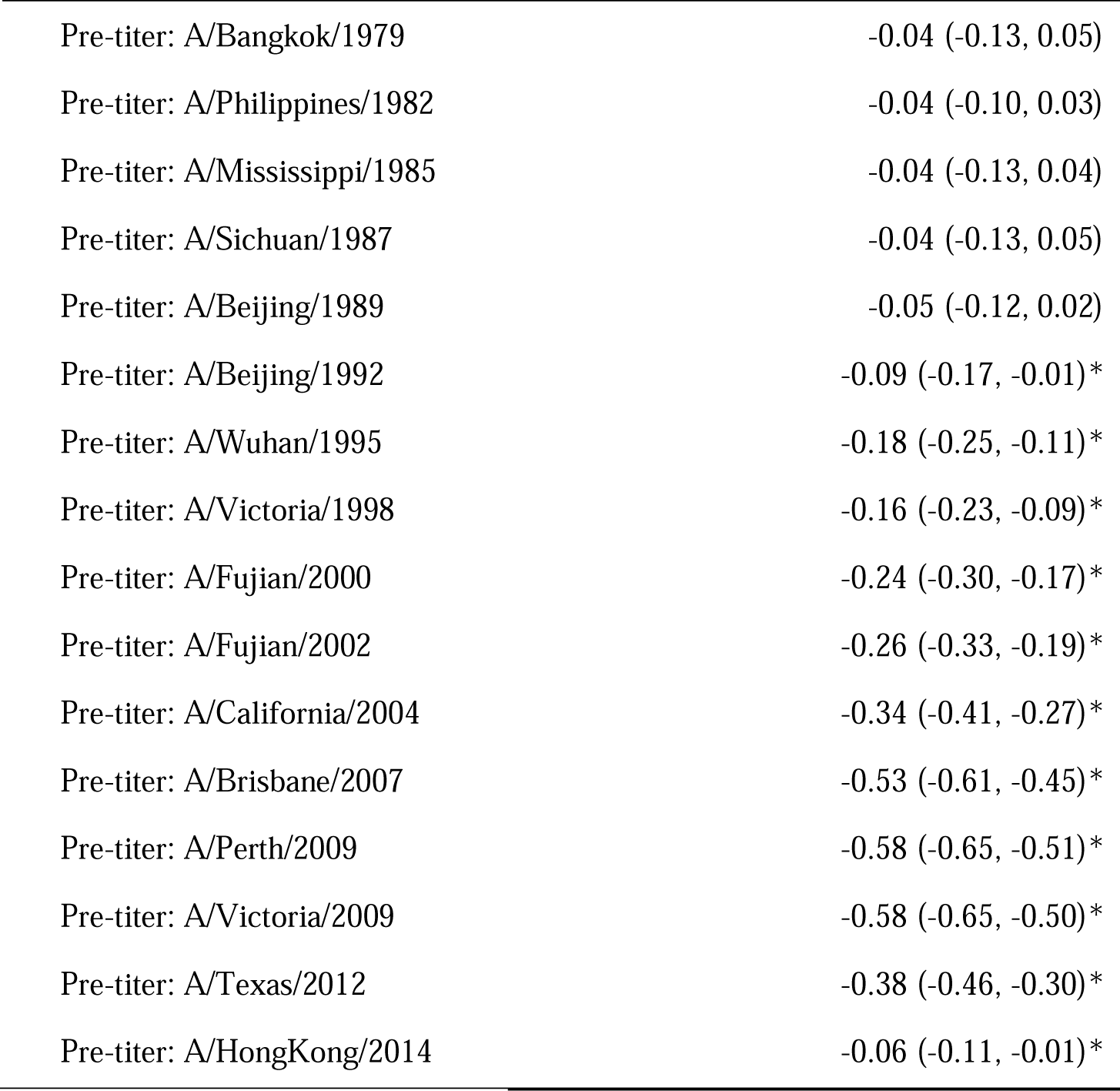
Association between pre-existing titer on changes of titers between two visits.

| Variable | Coefficient (95% confidence interval) |
| --- | --- |
| **Age** | 0.0030 (0.0019, 0.0040)* |
| **Strain** |  |
| A/HongKong/1968 | ref |
| X31 | -0.16 (-0.35, 0.03) |
| A/England/1972 | -0.13 (-0.32, 0.05) |
| A/Victoria/1975 | -0.01 (-0.18, 0.16) |
| A/Texas/1977 | 0.14 (-0.03, 0.32) |
| A/Bangkok/1979 | 0.00 (-0.16, 0.16) |
| A/Philippines/1982 | 0.01 (-0.17, 0.18) |
| A/Mississippi/1985 | -0.06 (-0.26, 0.15) |
| A/Sichuan/1987 | 0.20 (0.04, 0.37)* |
| A/Beijing/1989 | 0.04 (-0.12, 0.21) |
| A/Beijing/1992 | 0.02 (-0.18, 0.22) |
| A/Wuhan/1995 | 0.17 (0.00, 0.35)* |
| A/Victoria/1998 | 0.60 (0.41, 0.79)* |
| A/Fujian/2000 | 0.54 (0.35, 0.72)* |
| A/Fujian/2002 | 1.28 (1.06, 1.50)* |
| A/California/2004 | 1.10 (0.92, 1.28)* |
| A/Brisbane/2007 | 1.34 (1.17, 1.51)* |
| A/Perth/2009 | 1.52 (1.36, 1.68)* |
| A/Victoria/2009 | 2.35 (2.17, 2.52)* |
| A/Texas/2012 | 2.07 (1.90, 2.24)* |
| A/HongKong/2014 | 1.21 (1.04, 1.37)* |
| **Interact between pre-existing titer and strain** |  |
| Pre-titer: A/HongKong/1968 | -0.06 (-0.11, -0.01)* |
| Pre-titer: X31 | 0.04 (-0.03, 0.11) |
| Pre-titer: X31 | 0.02 (-0.05, 0.09) |
| Pre-titer: A/England/1972 | 0.01 (-0.07, 0.08) |
| Pre-titer: A/Victoria/1975 | -0.04 (-0.12, 0.03) |
| Pre-titer: A/Texas/1977 | -0.08 (-0.18, 0.01) |
| Pre-titer: A/Bangkok/1979 | -0.04 (-0.13, 0.05) |
| Pre-titer: A/Philippines/1982 | -0.04 (-0.10, 0.03) |
| Pre-titer: A/Mississippi/1985 | -0.04 (-0.13, 0.04) |
| Pre-titer: A/Sichuan/1987 | -0.04 (-0.13, 0.05) |
| Pre-titer: A/Beijing/1989 | -0.05 (-0.12, 0.02) |
| Pre-titer: A/Beijing/1992 | -0.09 (-0.17, -0.01)* |
| Pre-titer: A/Wuhan/1995 | -0.18 (-0.25, -0.11)* |
| Pre-titer: A/Victoria/1998 | -0.16 (-0.23, -0.09)* |
| Pre-titer: A/Fujian/2000 | -0.24 (-0.30, -0.17)* |
| Pre-titer: A/Fujian/2002 | -0.26 (-0.33, -0.19)* |
| Pre-titer: A/California/2004 | -0.34 (-0.41, -0.27)* |
| Pre-titer: A/Brisbane/2007 | -0.53 (-0.61, -0.45)* |
| Pre-titer: A/Perth/2009 | -0.58 (-0.65, -0.51)* |
| Pre-titer: A/Victoria/2009 | -0.58 (-0.65, -0.50)* |
| Pre-titer: A/Texas/2012 | -0.38 (-0.46, -0.30)* |
| Pre-titer: A/HongKong/2014 | -0.06 (-0.11, -0.01)* |

**Table S13.** Comparison of models of the association between pre-existing titer on changes of titers between two visits.

| **Model** | **Model terms** | **Adjusted R<sup>2</sup>** | **AIC** | **BIC** |
| --- | --- | --- | --- | --- |
| 1 | Pre-existing titer | 9.0 | 51211.7 | 51234.8 |
| 2 | Pre-existing titer + Age | 9.5 | 51134.5 | 51165.3 |
| 3 | Pre-existing titer + Strain | 22.8 | 48562.7 | 48739.8 |
| 4 | Pre-existing titer + Strain + Age | 23.2 | 48474.6 | 48659.4 |
| 5 | Pre-existing titer + Strain + Age + Pre-existing titer: Strain | 28.0 | 47441.5 | 47780.3 |

